# Blood biomarkers for Alzheimer disease across ethnoracial groups and healthcare settings: a systematic review and meta-analysis

**DOI:** 10.64898/2026.05.14.26353162

**Authors:** Xiaoxi Ma, Gurpreet Kaur Hansra, Tharusha Jayasena, John D Crawford, Anne Poljak, Perminder S Sachdev

## Abstract

Blood-based biomarkers could transform Alzheimer disease (AD) detection by enabling scalable, less invasive assessment of underlying pathology, yet their applicability across globally diverse populations remains uncertain. We systematically reviewed 168 publications comprising 139 independent cohorts from Asia, Europe, North America, South America, Africa and Oceania. Random-effects meta-analyses pooled log-transformed ratios of mean biomarker concentrations for AD versus cognitively unimpaired individuals, mild cognitive impairment versus cognitively unimpaired individuals, and amyloid-β PET-positive versus amyloid-β PET-negative individuals. p-tau217, p-tau181 and glial fibrillary acidic protein showed the largest and most consistent group differences. In clinically defined comparisons, p-tau181 separation in mild cognitive impairment was lower in predominantly Asian than White cohorts, whereas glial fibrillary acidic protein separation in AD was higher in predominantly Asian cohorts. No significant between-population differences were observed in amyloid-defined comparisons. These findings support leading blood biomarkers as globally relevant indicators of AD pathology, but rigorous harmonized validation is needed before thresholds can be translated into equitable clinical practice.

## Main

Alzheimer disease (AD) is the most common cause of dementia worldwide and an increasing public health challenge as populations age^1,2^. The emergence of disease-modifying therapies and biomarker-guided care has intensified the need for scalable tools that can identify AD pathology early and accurately^3-6^. Although cerebrospinal fluid and positron emission tomography (PET) biomarkers provide robust evidence of amyloid-β (Aβ) and tau pathology^2,7^, their use remains constrained by cost, invasiveness and limited availability, particularly outside specialist centers and high-income settings^4,8^. Blood-based biomarkers therefore have the potential to transform AD detection, triage and trial prescreening by enabling less invasive and more accessible assessment of underlying disease biology^3,4^.

Among the leading candidates, circulating phosphorylated tau species, glial fibrillary acidic protein (GFAP), neurofilament light chain (NfL) and amyloid-related measures have shown substantial promise for identifying AD pathology and distinguishing clinical stages of disease^3,4,9-15^. However, most biomarker development and validation studies have been conducted in relatively restricted populations, often in high-income countries and specialist memory-clinic cohorts^4,16^. This raises important questions about generalizability, threshold transportability and equitable implementation^4,16^. Reported differences in AD biomarkers across populations may reflect multiple overlapping influences, including age structure, vascular and renal comorbidity, assay platform, referral pathways, education and socioeconomic context, rather than differences in the biomarker-pathology relationship itself^4,16-23^. Yet current evidence remains fragmented, with limited representation of underrepresented populations and inconsistent use of biomarker-defined or pathology-confirmed reference standards^4,16,20-22^.

Here we performed a systematic review and meta-analysis of blood-based AD biomarkers across populations from Asia, Europe, North America, South America, Africa and Oceania. We compared biomarker differences between clinically defined groups and between amyloid-defined groups and examined whether study-level variation across populations was associated with ethnoracial composition, country income level, age, sex and diagnostic approach. We hypothesized that leading blood biomarkers would show a broadly consistent pathology-related signal across populations, but that apparent differences would be more evident in clinically defined comparisons than in pathology-defined analyses. By clarifying where blood biomarkers show cross-population consistency and where uncertainty remains, this study aims to inform their equitable global translation into research and clinical practice.

## Results

### Study selection and representation of populations

A total of 8167 records (6345 unique records) were identified for screening, of which 168 publications, comprising 139 independent studies or cohorts, were included in this systematic review and meta-analysis. Of these, 114 studies/cohorts contributed to clinically defined comparisons involving AD or mild cognitive impairment (MCI) versus cognitively unimpaired (CU) individuals, and 25 cohorts contributed to amyloid-β positron emission tomography (Aβ-PET)-defined comparisons between Aβ-positive and Aβ-negative individuals (Fig. 1). Included cohorts spanned Asia, Europe, North America, South America, Africa and Oceania, but representation was uneven. In clinically defined comparisons, most cohorts were predominantly Asian (k = 49) or White (k = 43), whereas Hispanic/Latino (k = 7), Black (k = 4), Indigenous (k = 1), Middle Eastern/North African (MENA, k = 3) and multi-ethnic cohorts (k = 7) were comparatively sparse. Asian cohorts were drawn predominantly from East Asia, with more limited representation from Southeast Asia and South Asia (Supplementary Table 1). Aβ-PET-defined analyses were even less diverse, consisting largely of White (k = 11) and Asian (k = 12) cohorts, with only two multi-ethnic cohorts (Supplementary Table 2). Blood-based biomarkers were measured across multiple assay platforms, most commonly single-molecule array (Simoa), alongside liquid chromatography–tandem mass spectrometry (LC-MS/MS), enzyme-linked immunosorbent assay (ELISA), Lumipulse, Meso Scale Discovery (MSD), Elecsys, immunomagnetic reduction (IMR), ELLA, Luminex xMAP and high-sensitivity chemiluminescence enzyme immunoassay (HISCL) (Supplementary Tables 1 and 2).

**Fig. 1.**
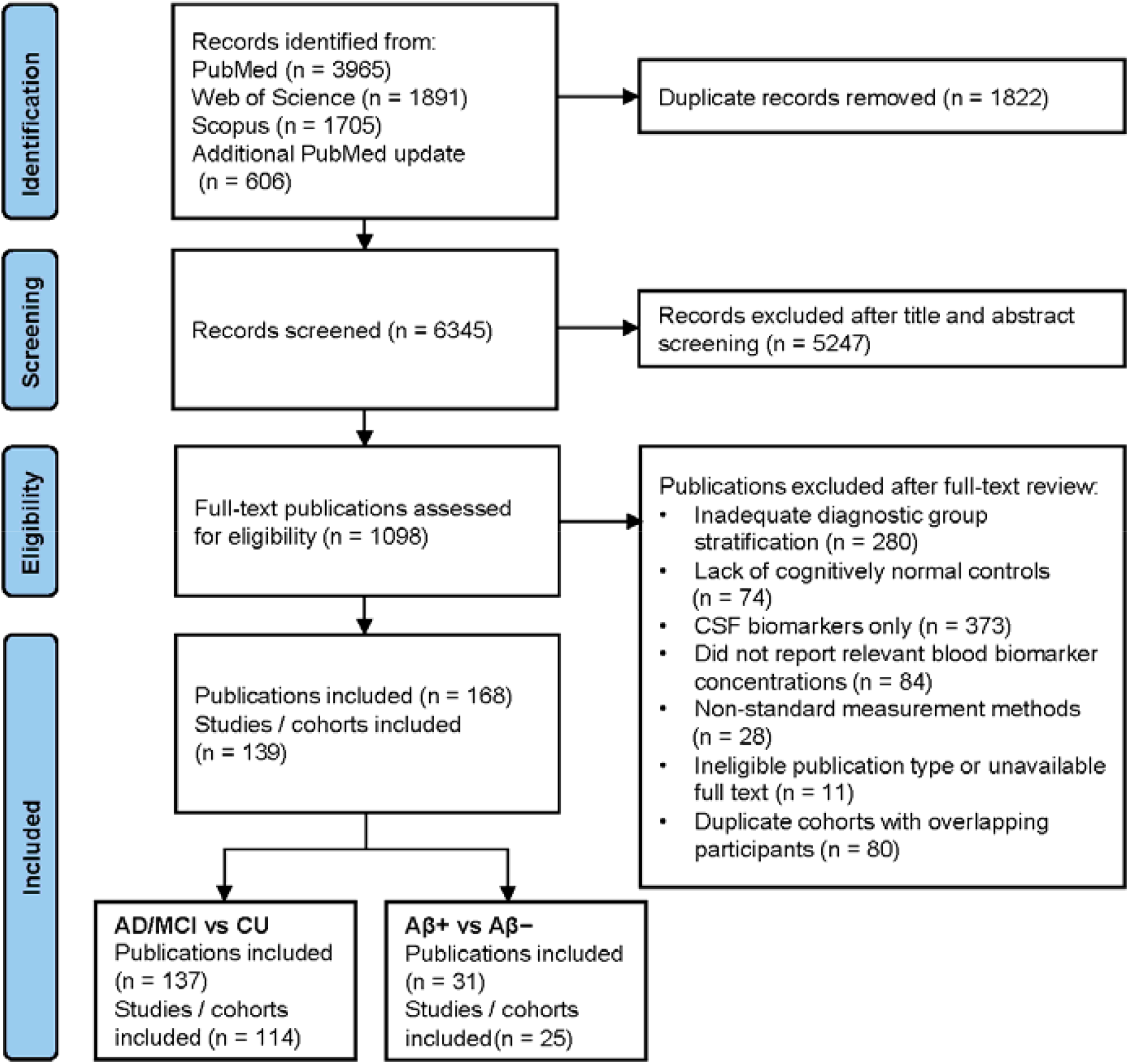
PRISMA flow diagram of study identification, screening and inclusion. Systematic searches of PubMed, Web of Science and Scopus identified studies published from database inception to 13 November 2024, followed by an updated PubMed search from 14 November 2024 to 8 August 2025. The diagram shows the numbers of records identified, screened, excluded and included in the systematic review and meta-analysis. Aβ+ and Aβ− denote amyloid-β positron emission tomography (Aβ-PET)-positive and Aβ-PET-negative groups, respectively. CSF, cerebrospinal fluid.

### Overall pooled biomarker differences in clinically defined comparisons

Across clinically defined comparisons, phosphorylated tau species and glial fibrillary acidic protein (GFAP) showed the largest average differences between disease groups, with neurofilament light chain (NfL) also showing consistent elevations (Table 1). In AD versus CU comparisons, the largest pooled ratios were observed for p-tau217 (ratio = 2.76, 95% CI 2.08–3.67), GFAP (1.94, 1.77–2.13) and p-tau181 (1.87, 1.72–2.02), followed by NfL (1.62, 1.53–1.73). In MCI versus CU comparisons, p-tau217 (1.95, 1.59–2.39), GFAP (1.47, 1.35–1.60) and p-tau181 (1.41, 1.31–1.53) again showed the largest pooled differences, whereas NfL showed a more modest but consistent elevation (1.27, 1.22–1.33) (Table 1). Amyloid-related measures were directionally consistent with AD biology, with reduced Aβ42/Aβ40 in both AD versus CU (0.87, 0.84–0.90) and MCI versus CU (0.91, 0.86–0.95), whereas Aβ42 was significantly reduced in AD versus CU (0.91, 0.84–0.97) but not in MCI versus CU (0.97, 0.91–1.04) (Table 1).

**Table 1.**
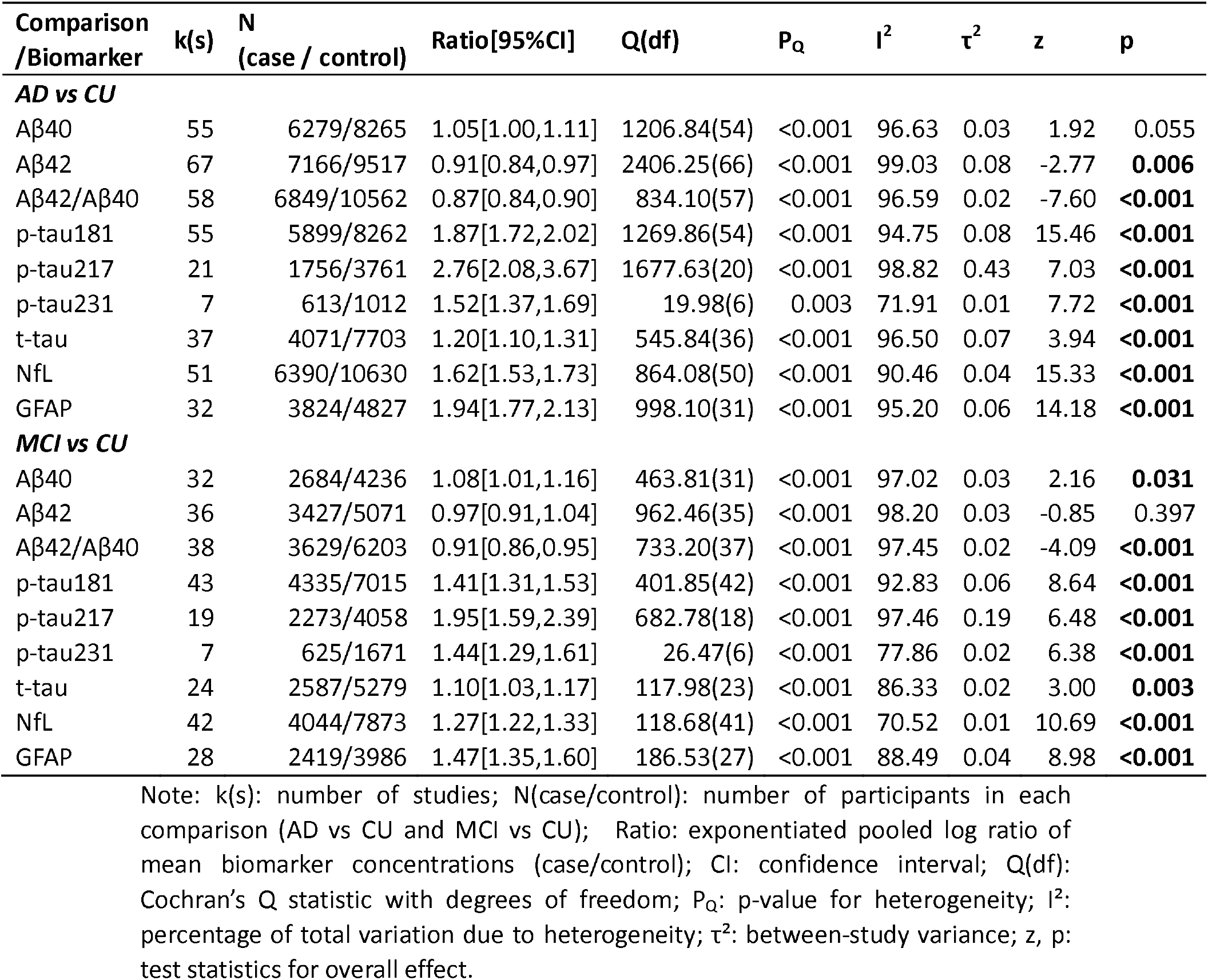
Meta-analysis of blood-based biomarkers comparing AD vs CU and MCI vs CU.

Between-study heterogeneity was substantial across nearly all biomarkers in clinically defined analyses (I^2^ > 70% for all biomarkers; all Cochran’s Q P < 0.001), indicating marked variability in study-level effect sizes across cohorts, assays and diagnostic settings (Table 1). Nonetheless, the overall pattern was robust in several sensitivity analyses. Exclusion of studies with Newcastle–Ottawa Scale scores <4 yielded materially similar pooled estimates (Supplementary Table 3). Analyses stratified by assay platform did not indicate that the principal findings were driven by a single platform (Supplementary Table 4). In studies including AD, MCI and CU groups, AD versus MCI ratios remained significantly elevated for p-tau217, p-tau181, NfL, GFAP, Aβ42/Aβ40, and Aβ42 whereas other biomarkers did not show significant differences (Supplementary Table 5). The principal findings were robust in sensitivity analyses restricted to plasma samples and excluding cohorts with inferred ethnoracial classification, with broadly similar pooled estimates observed in the clinically defined AD vs CU and MCI vs CU comparisons (Supplementary Table 6, 7). Leave-one-out analyses did not identify any single study that materially altered the direction of these overall effects, although a few borderline associations changed in statistical significance (Supplementary Table 8). Publication-bias diagnostics suggested possible small-study effects for selected biomarkers, but trim-and-fill adjustments resulted in only modest changes in pooled estimates and did not alter the overall direction of findings (Supplementary Fig. 1 and Supplementary Table 9).

### Study-level variation across populations in clinically defined comparisons

Across subgroup analyses, p-tau181, GFAP, NfL and Aβ42/Aβ40 showed broadly consistent directionality across the populations that could be evaluated, although precision varied substantially by subgroup size (Fig. 2 and Supplementary Table 10). P-tau217 significantly differentiated AD from CU in most populations, but the estimate in predominantly Asian cohorts did not reach conventional significance despite a large effect size (ratio = 2.07, 95% CI 0.99–4.32, P = 0.053), reflecting wide uncertainty (Fig. 2 and Supplementary Table 10). Significant differences for p-tau231 were identified only in White cohorts, for which data were available (ratio = 1.52, 95% CI 1.37–1.69 for AD versus CU), whereas data were insufficient for other populations (Supplementary Table 10). In Hispanic/Latino cohorts, most biomarkers did not show statistically detectable differences in MCI versus CU comparisons, and amyloid markers showed more variable subgroup patterns overall (Fig. 2 and Supplementary Table 10).

**Fig. 2.**
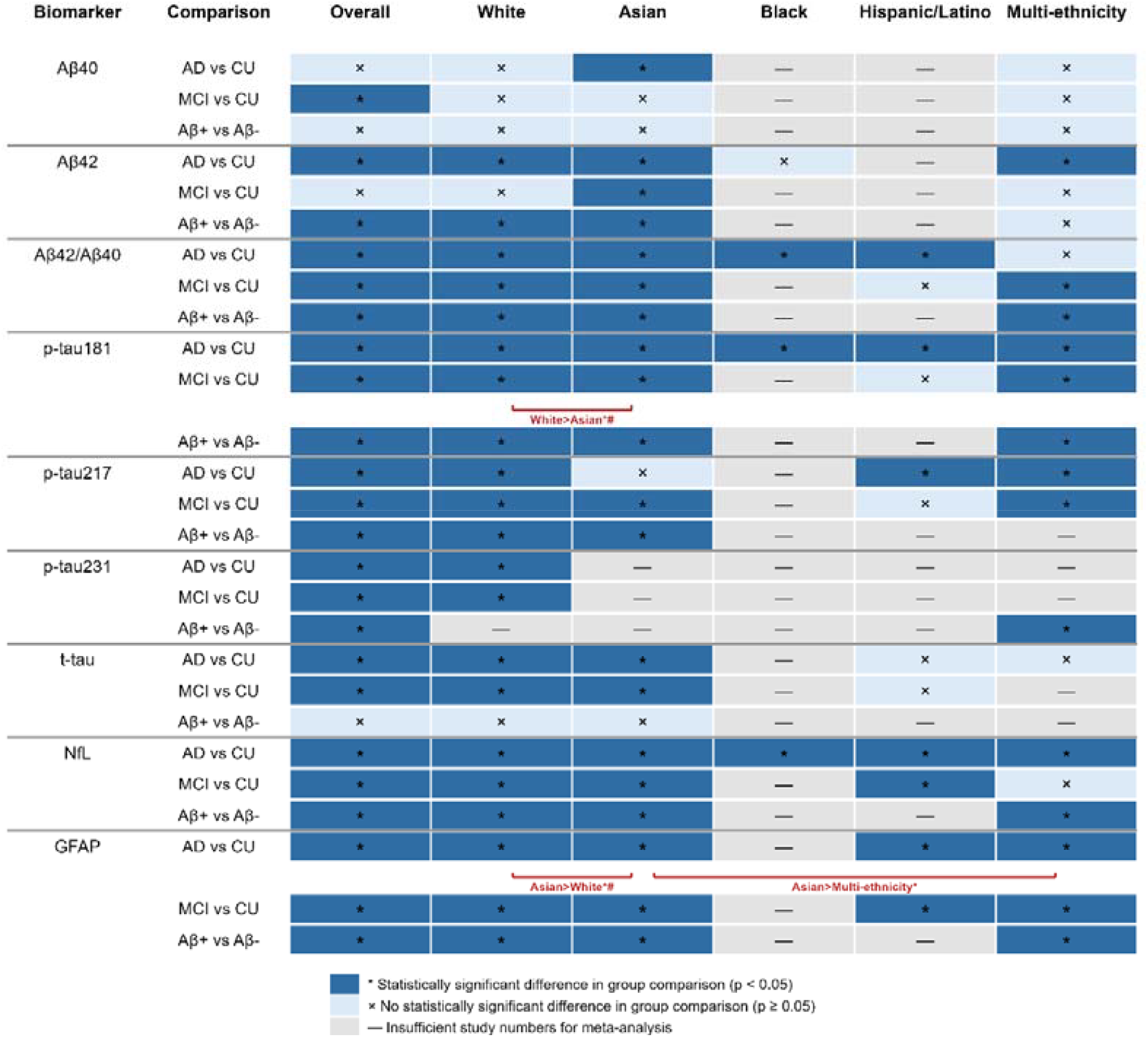
Summary of study-level blood biomarker differences across clinical and amyloid-defined comparisons. Cells indicate whether pooled random-effects meta-analyses identified statistically detectable differences in biomarker concentrations for AD versus cognitively unimpaired individuals (CU), mild cognitive impairment (MCI) versus CU, and Aβ-PET-positive versus Aβ-PET-negative individuals across populations defined by predominant cohort ethnoracial composition. Dark blue cells with check marks indicate P < 0.05; light blue cells with crosses indicate P ≥ 0.05; grey cells indicate insufficient data for meta-analysis (k < 2). Indigenous and MENA groups are not shown because data were insufficient. Red brackets indicate pairwise between-population differences identified in meta-regression; bracket direction indicates the population with the larger pooled ratio; *P < 0.05; #difference remained significant after adjustment for study-level age and sex where available.

Meta-regression identified two population contrasts that remained prominent in clinically defined analyses. The p-tau181 ratio for MCI versus CU was lower in predominantly Asian than in predominantly White cohorts (Asian: 1.27, 95% CI 1.15–1.41; White: 1.60, 95% CI 1.42–1.80; unadjusted P = 0.037; age- and sex-adjusted P = 0.03) (Figs. 2, 3 and Supplementary Table 10, 11). Conversely, the GFAP ratio for AD versus CU was higher in predominantly Asian than in predominantly White cohorts (Asian: 2.46, 95% CI 2.07–2.93; White: 1.89, 95% CI 1.71–2.10; unadjusted P = 0.017; age- and sex-adjusted P = 0.001) (Figs. 2, 4 and Supplementary Table 10, 11). No statistically detectable between-population differences were observed for the evaluable biomarkers in Hispanic/Latino or Black cohorts, although these analyses were based on limited numbers of studies and several biomarkers could not be assessed in these groups because of sparse data (Extended Data Figs. 1 –7 and Supplementary Table 11).

**Fig. 3.**
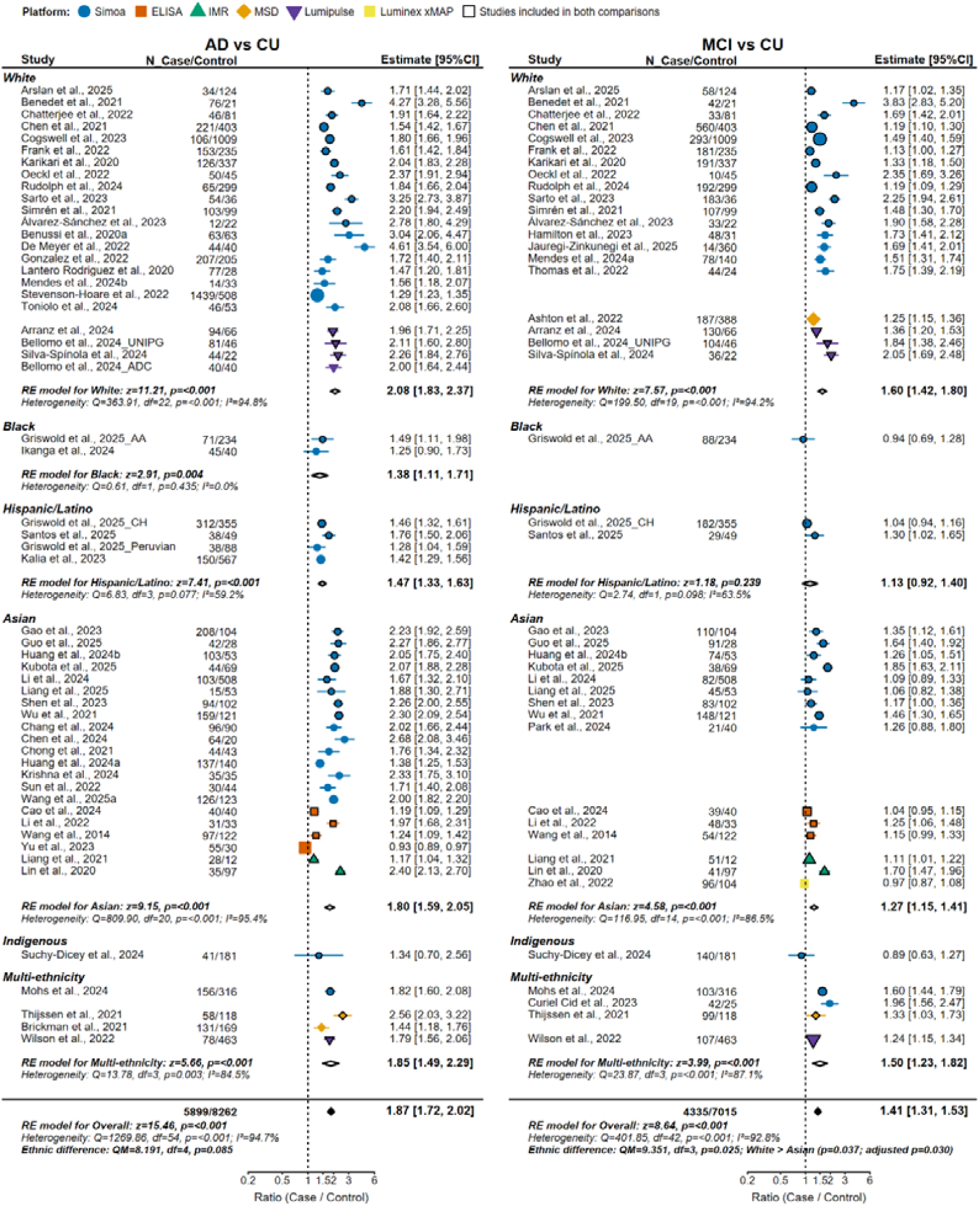
Forest plot of blood p-tau181 differences across populations. Forest plots show geometric mean ratios of blood p-tau181 concentrations for AD versus cognitively unimpaired individuals (left) and mild cognitive impairment versus cognitively unimpaired individuals (right), stratified by predominant cohort ethnoracial composition. Points indicate study-specific estimates and 95% confidence intervals (CIs); colours and shapes denote assay platforms; markers outlined in black indicate studies contributing to both comparisons. Hollow diamonds indicate pooled random-effects estimates within each population subgroup; filled diamonds indicate the overall pooled effect. For each subgroup and for the overall analysis, Cochran’s Q statistic, degrees of freedom (df), I^2^ and the test of the pooled effect are shown. The between-subgroup heterogeneity statistic (Q_M) is shown for the overall model. For significant pairwise subgroup contrasts, unadjusted and age- and sex-adjusted P values are reported where available.

**Fig. 4.**
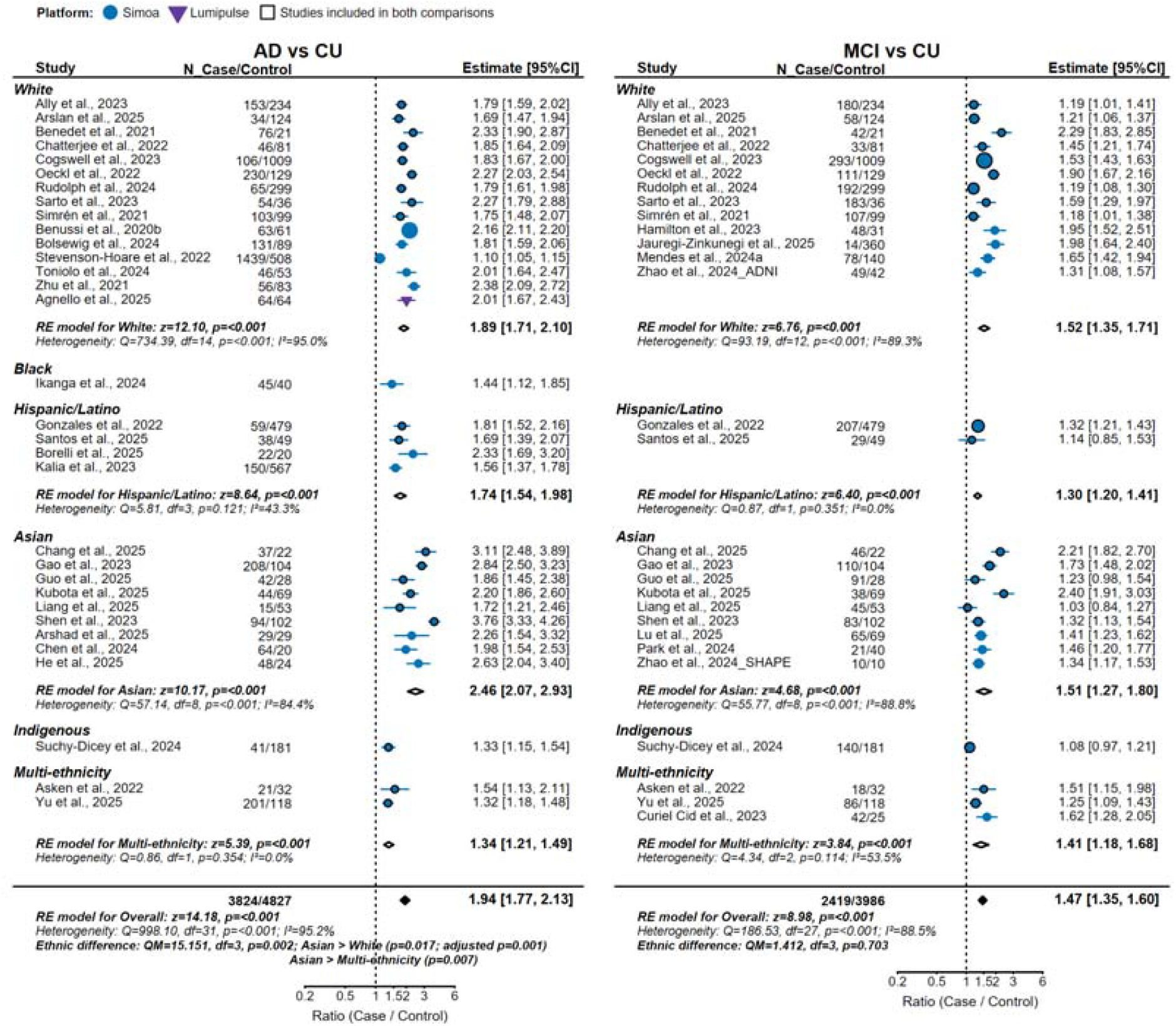
Forest plot of blood GFAP differences across populations. Forest plots show geometric mean ratios of blood glial fibrillary acidic protein (GFAP) concentrations for AD versus cognitively unimpaired individuals (left) and mild cognitive impairment versus cognitively unimpaired individuals (right), stratified by predominant cohort ethnoracial composition. Points indicate study-specific estimates and 95% confidence intervals (CIs); colours and shapes denote assay platforms; markers outlined in black indicate studies contributing to both comparisons. Hollow diamonds indicate pooled random-effects estimates within each population subgroup; filled diamonds indicate the overall pooled effect. For each subgroup and for the overall analysis, Cochran’s Q statistic, degrees of freedom (df), I^2^ and the test of the pooled effect are shown. The between-subgroup heterogeneity statistic (Q_M) is shown for the overall model. For significant pairwise subgroup contrasts, unadjusted and age- and sex-adjusted P values are reported where available.

The direction of the Asian–White differences for p-tau181 and GFAP was similar in analyses restricted to studies including AD, MCI and CU groups, whereas no significant between-population differences were observed for AD versus MCI comparisons (Supplementary Table 5). Platform-restricted analyses within Simoa also yielded results broadly consistent with the primary analyses where more than one population group was represented (Supplementary Table 4). Restriction to plasma samples and exclusion of cohorts with inferred ethnoracial classification yielded similar results across White and Asian cohorts for p-tau181, GFAP, NfL and Aβ42/Aβ40, although precision was lower in smaller population groups (Supplementary Table 6, 7). Leave-one-out analyses likewise indicated that most subgroup findings were stable, with changes in significance largely confined to borderline associations (Supplementary Table 8). Taken together, these findings indicate that variation across populations was more evident in clinically defined comparisons than in the overall pooled biomarker ranking itself.

### Amyloid PET-defined comparisons show broad cross-population consistency

In Aβ-PET-defined analyses, p-tau217, p-tau181 and GFAP again showed the largest average group differences (Table 2). The pooled Aβ-positive versus Aβ-negative ratio was 2.98 (95% CI 2.22–4.00) for p-tau217, 1.67 (1.51–1.86) for p-tau181 and 1.56 (1.46–1.68) for GFAP. Significant but smaller elevations were also observed for p-tau231 (1.37, 1.11–1.70) and NfL (1.23, 1.16–1.30). In contrast, Aβ42 (0.89, 0.86–0.92) and Aβ42/Aβ40 (0.86, 0.84–0.89) were significantly reduced in Aβ-positive individuals, whereas Aβ40 and t-tau did not differ significantly by amyloid PET status (Table 2). Egger’s tests did not indicate clear small-study effects in the Aβ-PET-defined analyses, and Trim-and-Fill adjustment had minimal impact on the pooled estimates (Supplementary Fig. 1 and Supplementary Table 9).

**Table 2.**
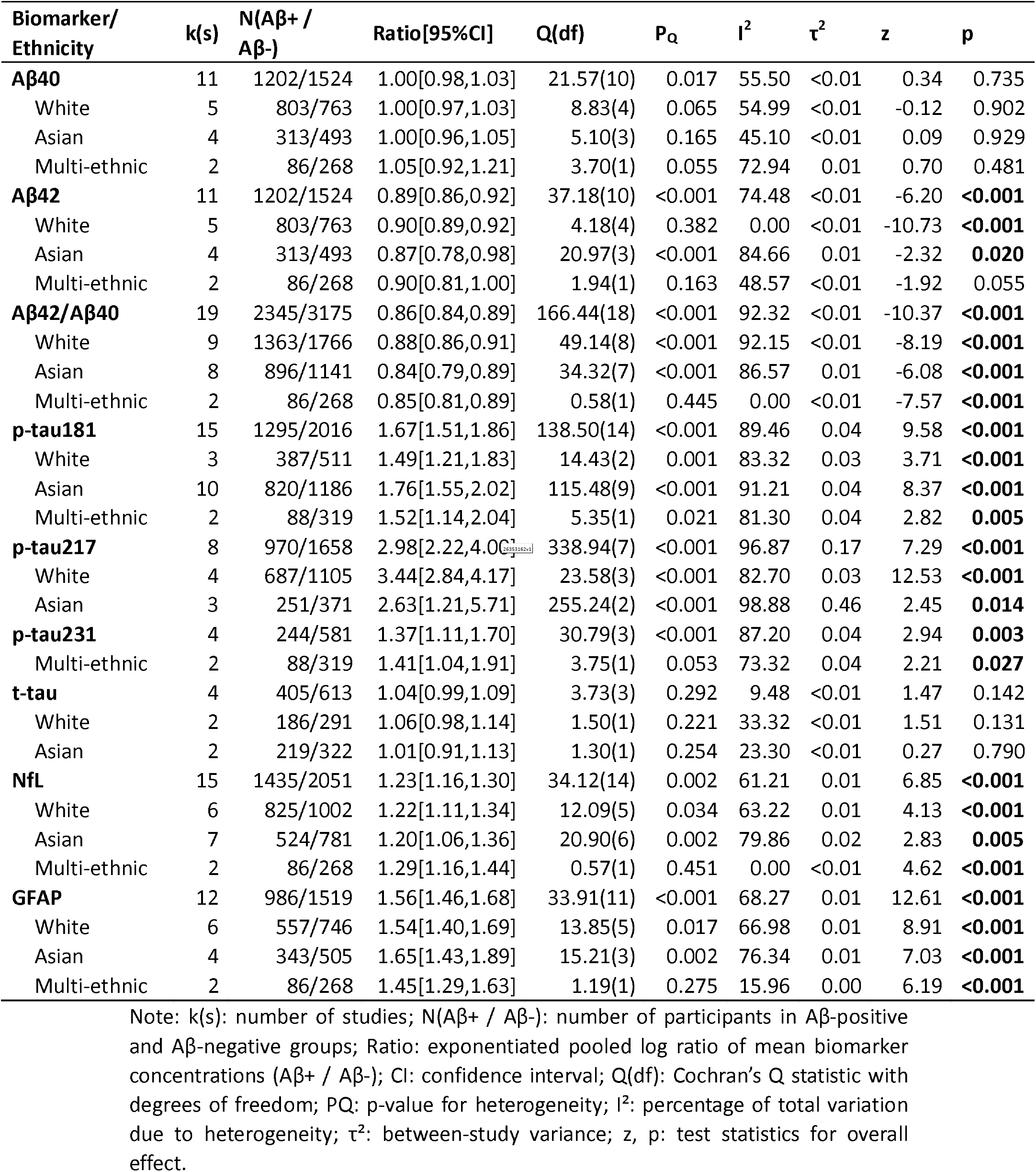
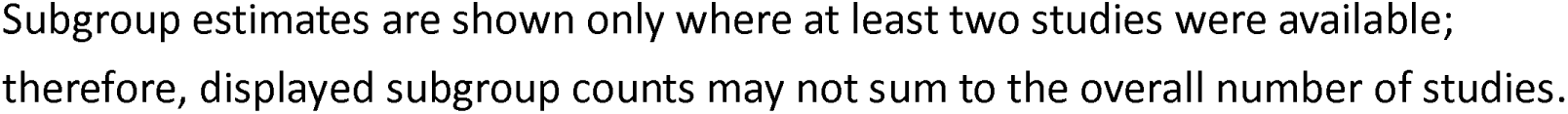
Meta-analysis of blood-based biomarker ratios in Aβ+ vs Aβ− comparisons, overall and by predominant cohort ethnoracial composition.

| **Biomarker/**<br>**Ethnicity** | **k(s)** | **N(Aβ+ /**<br>**Aβ-)** | **Ratio[95%CI]** | **Q(df)** | **P<sub>Q</sub>** | **I<sup>2</sup>** | **$\tau^2$** | **z** | **p** |
| --- | --- | --- | --- | --- | --- | --- | --- | --- | --- |
| **Aβ40** | 11 | 1202/1524 | 1.00[0.98,1.03] | 21.57(10) | 0.017 | 55.50 | <0.01 | 0.34 | 0.735 |
| White | 5 | 803/763 | 1.00[0.97,1.03] | 8.83(4) | 0.065 | 54.99 | <0.01 | -0.12 | 0.902 |
| Asian | 4 | 313/493 | 1.00[0.96,1.05] | 5.10(3) | 0.165 | 45.10 | <0.01 | 0.09 | 0.929 |
| Multi-ethnic | 2 | 86/268 | 1.05[0.92,1.21] | 3.70(1) | 0.055 | 72.94 | 0.01 | 0.70 | 0.481 |
| **Aβ42** | 11 | 1202/1524 | 0.89[0.86,0.92] | 37.18(10) | <0.001 | 74.48 | <0.01 | -6.20 | **<0.001** |
| White | 5 | 803/763 | 0.90[0.89,0.92] | 4.18(4) | 0.382 | 0.00 | <0.01 | -10.73 | **<0.001** |
| Asian | 4 | 313/493 | 0.87[0.78,0.98] | 20.97(3) | <0.001 | 84.66 | 0.01 | -2.32 | **0.020** |
| Multi-ethnic | 2 | 86/268 | 0.90[0.81,1.00] | 1.94(1) | 0.163 | 48.57 | <0.01 | -1.92 | 0.055 |
| **Aβ42/Aβ40** | 19 | 2345/3175 | 0.86[0.84,0.89] | 166.44(18) | <0.001 | 92.32 | <0.01 | -10.37 | **<0.001** |
| White | 9 | 1363/1766 | 0.88[0.86,0.91] | 49.14(8) | <0.001 | 92.15 | <0.01 | -8.19 | **<0.001** |
| Asian | 8 | 896/1141 | 0.84[0.79,0.89] | 34.32(7) | <0.001 | 86.57 | 0.01 | -6.08 | **<0.001** |
| Multi-ethnic | 2 | 86/268 | 0.85[0.81,0.89] | 0.58(1) | 0.445 | 0.00 | <0.01 | -7.57 | **<0.001** |
| **p-tau181** | 15 | 1295/2016 | 1.67[1.51,1.86] | 138.50(14) | <0.001 | 89.46 | 0.04 | 9.58 | **<0.001** |
| White | 3 | 387/511 | 1.49[1.21,1.83] | 14.43(2) | 0.001 | 83.32 | 0.03 | 3.71 | **<0.001** |
| Asian | 10 | 820/1186 | 1.76[1.55,2.02] | 115.48(9) | <0.001 | 91.21 | 0.04 | 8.37 | **<0.001** |
| Multi-ethnic | 2 | 88/319 | 1.52[1.14,2.04] | 5.35(1) | 0.021 | 81.30 | 0.04 | 2.82 | **0.005** |
| **p-tau217** | 8 | 970/1658 | 2.98[2.22,4.00] | 338.94(7) | <0.001 | 96.87 | 0.17 | 7.29 | **<0.001** |
| White | 4 | 687/1105 | 3.44[2.84,4.17] | 23.58(3) | <0.001 | 82.70 | 0.03 | 12.53 | **<0.001** |
| Asian | 3 | 251/371 | 2.63[1.21,5.71] | 255.24(2) | <0.001 | 98.88 | 0.46 | 2.45 | **0.014** |
| **p-tau231** | 4 | 244/581 | 1.37[1.11,1.70] | 30.79(3) | <0.001 | 87.20 | 0.04 | 2.94 | **0.003** |
| Multi-ethnic | 2 | 88/319 | 1.41[1.04,1.91] | 3.75(1) | 0.053 | 73.32 | 0.04 | 2.21 | **0.027** |
| **t-tau** | 4 | 405/613 | 1.04[0.99,1.09] | 3.73(3) | 0.292 | 9.48 | <0.01 | 1.47 | 0.142 |
| White | 2 | 186/291 | 1.06[0.98,1.14] | 1.50(1) | 0.221 | 33.32 | <0.01 | 1.51 | 0.131 |
| Asian | 2 | 219/322 | 1.01[0.91,1.13] | 1.30(1) | 0.254 | 23.30 | <0.01 | 0.27 | 0.790 |
| **NfL** | 15 | 1435/2051 | 1.23[1.16,1.30] | 34.12(14) | 0.002 | 61.21 | 0.01 | 6.85 | **<0.001** |
| White | 6 | 825/1002 | 1.22[1.11,1.34] | 12.09(5) | 0.034 | 63.22 | 0.01 | 4.13 | **<0.001** |
| Asian | 7 | 524/781 | 1.20[1.06,1.36] | 20.90(6) | 0.002 | 79.86 | 0.02 | 2.83 | **0.005** |
| Multi-ethnic | 2 | 86/268 | 1.29[1.16,1.44] | 0.57(1) | 0.451 | 0.00 | <0.01 | 4.62 | **<0.001** |
| **GFAP** | 12 | 986/1519 | 1.56[1.46,1.68] | 33.91(11) | <0.001 | 68.27 | 0.01 | 12.61 | **<0.001** |
| White | 6 | 557/746 | 1.54[1.40,1.69] | 13.85(5) | 0.017 | 66.98 | 0.01 | 8.91 | **<0.001** |
| Asian | 4 | 343/505 | 1.65[1.43,1.89] | 15.21(3) | 0.002 | 76.34 | 0.01 | 7.03 | **<0.001** |
| Multi-ethnic | 2 | 86/268 | 1.45[1.29,1.63] | 1.19(1) | 0.275 | 15.96 | 0.00 | 6.19 | **<0.001** |

Note: k(s): number of studies; N(Aβ + / Aβ -): number of participants in Aβ-positive and Aβ-negative groups; Ratio: exponentiated pooled log ratio of mean biomarker concentrations (Aβ + / Aβ -); CI: confidence interval; Q(df): Cochran's Q statistic with degrees of freedom; P<sub>Q</sub>: p-value for heterogeneity; I<sup>2</sup>: percentage of total variation due to heterogeneity; $\tau^2$ : between-study variance; z, p: test statistics for overall effect.
Subgroup estimates are shown only where at least two studies were available; therefore, displayed subgroup counts may not sum to the overall number of studies.

As in the clinically defined analyses, heterogeneity remained moderate to substantial for several biomarkers in Aβ-PET-defined comparisons, particularly for phosphorylated tau markers and Aβ42/Aβ40 (Table 2). However, in contrast to clinically defined comparisons, no statistically significant between-population differences were detected across the biomarkers that could be evaluated, and ethnicity-specific subgroup estimates were directionally consistent with the overall pooled effects (Table 2, Supplementary Table 11, and Extended Data Figs. 8–10). These analyses were dominated by White and Asian cohorts, with very limited representation from multi-ethnic populations, so the absence of detectable differences should be interpreted in the context of a restricted evidence base (Supplementary Table 2). Even so, the relative stability of Aβ-PET-defined findings across the populations most studied suggests that leading blood biomarkers track underlying amyloid pathology more consistently than clinically defined group status. Sensitivity analyses of the Aβ+ vs Aβ− comparisons were broadly consistent with the main results, both overall and across the populations that could be evaluated. Platform-restricted analyses suggested that the principal signals were not driven by a single assay platform, and similar overall and subgroup patterns were seen after restriction to plasma-only studies and to studies with explicit or predominant ethnoracial classification (Supplementary Table 4, 6, 7). Leave-one-out analyses likewise supported the stability of the main results, with changes in statistical significance largely limited to smaller or borderline subgroup estimates (Supplementary Table 8).

### Exploratory analyses of sources of heterogeneity

We next performed exploratory interaction and subgroup analyses to assess whether study-level variation in clinically defined comparisons was associated with demographic, diagnostic and contextual factors, focusing primarily on contrasts between Asian and White cohorts (Fig. 5 and Supplementary Tables 12–22). Age was associated with the Asian–White difference in AD versus CU GFAP ratios (β = −0.47, 95% CI −0.72 to −0.22, P = 0.001), with the between-population difference becoming less pronounced in older study populations (Fig. 5A and Supplementary Tables 12). Similar age-related interaction patterns were also observed in AD versus CU contrasts for GFAP involving Hispanic/Latino, White and Asian cohorts, and for Aβ40 involving Asian, White and multi-ethnic cohorts (Supplementary Tables 12). By contrast, sex did not show significant interaction effects for the principal biomarker comparisons evaluated (Fig. 5B and Supplementary Tables 13).

**Fig. 5.**
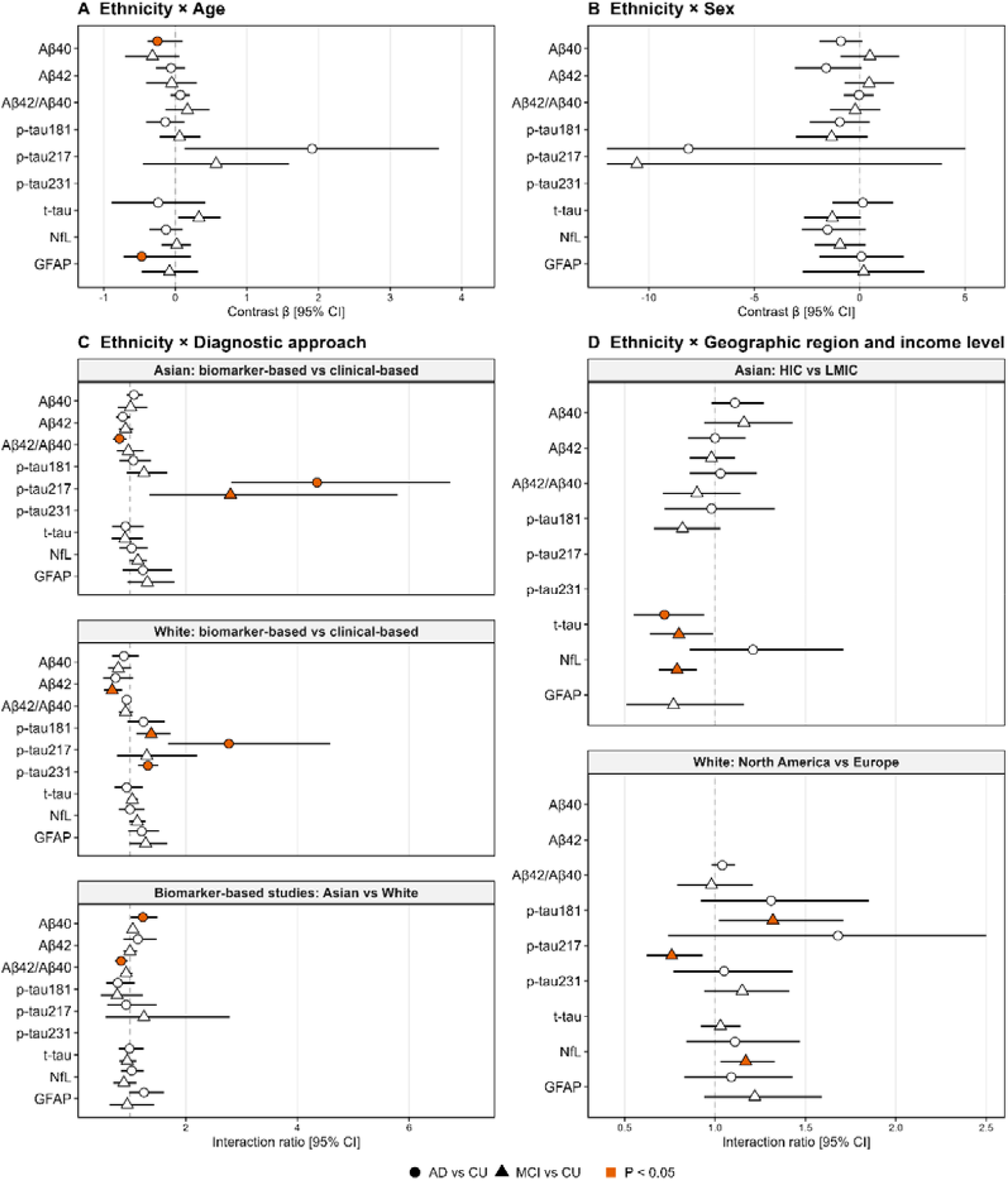
Exploratory interaction analyses of study-level differences between predominantly Asian and predominantly White cohorts. Forest plots show interaction effects for age (a), sex (b), diagnostic approach (c), and geographic region or country income level (d) on biomarker ratios in AD versus cognitively unimpaired individuals and mild cognitive impairment versus cognitively unimpaired individuals. Effect sizes are shown as contrast β values for continuous moderators (age and sex) or interaction ratios for categorical moderators (diagnostic approach, region and income level), each with 95% confidence intervals (CIs). Circles indicate AD versus cognitively unimpaired comparisons; triangles indicate mild cognitive impairment versus cognitively unimpaired comparisons; filled symbols indicate P < 0.05; open symbols indicate P ≥ 0.05; dashed vertical lines indicate the null value (β = 0 or ratio = 1). HIC, high-income country; LMIC, low- and middle-income country.

Diagnostic approach was also associated with study-level biomarker contrasts. In Asian cohorts, significant subgroup differences by diagnostic approach were observed for Aβ42/Aβ40 and p-tau217, whereas in White cohorts they were observed for Aβ42, p-tau181, p-tau217 and p-tau231 in specific comparisons (Fig. 5C and Supplementary Tables 14 and 15). Where significant, biomarker-based studies generally showed more pronounced case–control differences than clinical-based studies. When analyses were restricted to biomarker-based studies, the Asian–White differences observed for p-tau181 and GFAP were no longer statistically significant, whereas significant differences emerged for Aβ40 (P = 0.041) and Aβ42/Aβ40 (P = 0.009) in AD versus CU comparisons (Fig. 5C and Supplementary Table 16). These findings further suggest that variation in case definition contributes to the population differences observed in clinically defined analyses.

Contextual analyses also indicated variation by country income level and region, although these findings should be considered exploratory. Within Asian cohorts, country income level was associated with AD versus CU (P = 0.017) and MCI versus CU (P = 0.039) ratios of t-tau, and MCI versus CU ratios of NfL (P < 0.001), with larger effect sizes observed in high-income than low- and middle-income settings; these patterns were largely driven by East Asian cohorts (Fig. 5D and Supplementary Tables 17 and 18). Within White cohorts, AD versus CU biomarker ratios were broadly comparable between North American and European studies, although regional differences were observed for p-tau181, p-tau217 and NfL in MCI versus CU comparisons (Supplementary Table 19). Because data from Hispanic/Latino and Black cohorts were limited, regional analyses in these groups were descriptive only (Supplementary Tables 20 and 21). In North American analyses comparing predominantly White and multi-ethnic cohorts, significant interactions were observed for GFAP and NfL in AD versus CU comparisons and for Aβ42/Aβ40 in MCI versus CU comparisons, whereas most other contrasts were similar across groups (Supplementary Table 22).

## Discussion

Blood-based biomarkers are moving rapidly from research tools toward practical instruments for triage, trial enrichment and, increasingly, clinical decision-making in AD. In this systematic review and meta-analysis of 168 publications comprising 139 independent cohorts, p-tau217, p-tau181 and GFAP showed the largest and most consistent study-level differences across AD-related group comparisons. Across clinically defined analyses, p-tau217 showed the greatest average separation between AD and cognitively unimpaired individuals, with p-tau181 and GFAP also showing robust differences; in amyloid PET-defined comparisons, the same markers again showed the strongest signal. By contrast, amyloid-related measures and NfL were directionally informative but more variable across studies. These findings are concordant with the broader literature positioning phosphorylated tau species, particularly p-tau217 and p-tau181, as the most informative blood-based markers of AD pathology, with GFAP emerging as an important complementary marker linked to early amyloid-associated astroglial responses^4,9,10,13,14,24-28^.

The principal advance of this study lies not only in the overall biomarker ranking, but in the contrast between clinically defined and pathology-defined analyses. Study-level variation across populations was more apparent when groups were defined by clinical diagnosis than when they were defined by amyloid PET status. In clinically defined comparisons, the p-tau181 ratio for MCI versus cognitively unimpaired individuals was lower in predominantly Asian than predominantly White cohorts, whereas the GFAP ratio for AD versus cognitively unimpaired individuals was higher in predominantly Asian cohorts. However, no statistically detectable between-population differences were observed in amyloid PET-defined analyses for the biomarkers that could be evaluated. This pattern suggests that the relationship between leading blood biomarkers and underlying amyloid pathology is broadly consistent across the populations most extensively studied, whereas variation in clinically defined group separation is more likely to reflect differences in case definition, referral pathways, disease stage at ascertainment and broader study context. That interpretation is consistent with prior work showing that apparent ethnoracial differences in CSF, PET and blood biomarkers are often reduced after accounting for socioeconomic, clinical and methodological factors^17-22^.

This distinction has direct translational relevance. A blood biomarker may retain stable pathology-related signal across populations while showing variable separation between clinically defined groups if those groups are assembled differently across settings. Clinical diagnoses of AD and MCI are shaped by access to specialist care, the use of confirmatory biomarkers, education and language, competing neuropathologies, and the burden of vascular, renal and other systemic comorbidities^3,4,16,20-23,29,30^. These factors can alter the case mix even when the underlying biomarker-pathology relationship is preserved. Our findings therefore argue against equating clinically defined group differences with biological non-equivalence. For implementation, the more relevant signal is the relative stability of pathology-linked biomarker differences; the variability observed in clinically defined comparisons instead points to the need for calibrated diagnostic pathways and careful local validation.

The overall biomarker hierarchy observed here aligns with the current direction of the field. Plasma p-tau217 showed the largest average differences across both clinically defined and amyloid-defined analyses, reinforcing its position as the leading blood-based marker of AD-related pathological change^10,24-27,31,32^. P-tau181 also showed strong and reproducible differences across comparisons and populations, supporting its continued importance where p-tau217 is unavailable or less standardized^9,12,24,28^. GFAP emerged as a strong complementary marker in both clinical and amyloid-defined comparisons, consistent with evidence that plasma GFAP rises early with amyloid pathology and may capture astroglial activation along the AD continuum^13,14^. NfL remained informative as an index of neuroaxonal injury, but its broader sensitivity to non-AD neurodegeneration probably reduces disease specificity in heterogeneous clinical cohorts^4,15,33^. Amyloid-related measures, particularly Aβ42/Aβ40, were directionally consistent with AD biology but more susceptible to variability across studies, likely reflecting both biological and analytical influences^34-37^.

The observed Asian-White contrasts should, however, be interpreted cautiously. First, these analyses were conducted at the study level and therefore cannot distinguish participant-level biological differences from ecological differences in recruitment, ascertainment, comorbidity structure or health-care setting. Second, the Asian category encompassed highly heterogeneous populations and was dominated by East Asian cohorts, with more limited representation from South and Southeast Asia. Third, several exploratory analyses suggested that some clinically defined differences were attenuated or no longer statistically apparent after stratification by diagnostic approach and other study-level characteristics. Accordingly, the most defensible interpretation is not that the present data demonstrate ethnicity-specific biomarker-pathology relationships, but that some biomarkers show different degrees of clinical group separation across populations because the clinical groups themselves are not defined or ascertained in equivalent ways across settings. This interpretation is also consistent with recent multi-ethnic cohort studies showing comparable biomarker-pathology relationships across groups despite differences in biomarker distributions or optimal prescreen thresholds^20-22,31^.

The exploratory moderator analyses further support a contextual interpretation. Age was associated with variation in GFAP-related contrasts across populations, which is biologically plausible given the sensitivity of GFAP to ageing, astroglial activation and mixed brain injury^13,14,38,39^. Diagnostic approach also appeared to matter: biomarker-confirmed studies generally showed larger effect sizes for some markers, and some between-population differences seen in clinically defined analyses were no longer sustained in biomarker-based subsets. Variation by country income level and geographic region was also observed for selected markers, including t-tau and NfL within Asian cohorts, although these analyses should be regarded as exploratory. Such patterns may reflect differences in vascular and renal disease burden, stage at presentation, access to specialist assessment, case selection and health-system organization^23,40,41^. Because these moderators were available only at the study level and often clustered together, their independent contributions cannot be resolved here. Nonetheless, the data underscores a key principle for translation: interpretation of blood biomarkers is inseparable from the context in which patients are identified and classified.

A further barrier to implementation is technical heterogeneity. Absolute concentrations of blood biomarkers can vary across assay platforms, sample matrices and pre-analytical workflows, complicating the transferability of thresholds across studies and health-care systems^36,42-44^. By pooling ratios of means rather than absolute concentrations, we sought to reduce calibration-dependent variation and focus on relative group separation. Even with this approach, heterogeneity remained substantial across most biomarkers, indicating that assay characteristics, laboratory implementation and sample handling probably contributed materially to between-study variability. This distinction is important for practice: a biomarker can show broadly consistent pathology-related signal across populations, yet still require harmonized calibration, reference materials and setting-specific validation before it can be deployed confidently in routine care. In other words, biological generalizability does not automatically imply threshold portability.

These findings therefore support a clinically meaningful but appropriately bounded conclusion. The data are encouraging for the global development of blood-based AD biomarkers, particularly p-tau217, p-tau181 and GFAP, as scalable tools for pathology-oriented assessment, triage and research enrichment^3,45^. At the same time, the present evidence does not justify population-specific thresholds, nor does it support uncritical transfer of thresholds or screening algorithms from one health-care system to another^43,44^. For clinicians, the message is not that blood biomarkers fail across diverse populations, but that implementation should proceed through harmonized validation rather than assumption^43-45^. For researchers and trialists, the results reinforce the value of pathology-confirmed, globally diverse cohorts and the need to test whether calibration, discrimination and clinical utility remain stable across settings before biomarker-driven pathways are generalized^16,43,44^.

Several limitations frame these conclusions. Most importantly, this study synthesized study-level contrasts in biomarker concentrations rather than individual-level diagnostic accuracy and therefore cannot determine sensitivity, specificity, calibration, optimal thresholds or real-world clinical utility. Population categories were assigned at the cohort level and reflect predominant study composition rather than harmonized participant-level measures of self-identified ethnicity or genetic ancestry; the analyses are therefore vulnerable to ecological misclassification and cannot disentangle ethnicity from geography, socioeconomic context or health-system differences. Between-study heterogeneity was high across most biomarkers, limiting the transportability of pooled estimates. Many clinically defined cohorts lacked pathology confirmation, increasing the risk of diagnostic misclassification. Finally, the evidence base remained heavily weighted toward Asian and White cohorts, with sparse data from Black, Hispanic/Latino, Indigenous, African, South Asian and several low- and middle-income populations, particularly in amyloid PET-defined analyses. Apparent similarities or differences in underrepresented groups should therefore be interpreted with caution rather than as evidence of equivalence or absence of effect.

In summary, p-tau217, p-tau181 and GFAP showed the most robust blood-based signal across AD-related comparisons, and leading biomarkers appeared to track amyloid pathology broadly consistently across the populations most extensively studied. Apparent between-population differences were more evident in clinically defined analyses than in amyloid-defined comparisons, consistent with an important contribution from diagnostic and contextual heterogeneity rather than clear evidence for ethnicity-specific biomarker biology. The immediate implication is high-impact and practical: blood biomarkers are now credible candidates for broader global AD assessment, but the field must move beyond proof-of-concept toward harmonized validation in the populations and health systems that have so far been least represented. The next phase should define whether common thresholds, calibration strategies and clinical pathways can deliver equitable biomarker-based care at scale. Until then, the strongest conclusion is one of cautious confidence: these biomarkers are ready for global development, but not yet for indiscriminate global deployment.

## Methods

### Study design and reporting

This systematic review and meta-analysis was prospectively registered in PROSPERO (CRD420251031990) and reported in accordance with the Preferred Reporting Items for Systematic Reviews and Meta-Analyses (PRISMA) 2020 statement^46^.

### Information sources and search strategy

Two authors (X.M. and G.H.) independently searched PubMed, Web of Science and Scopus from database inception to 13 November 2024. An updated search was subsequently performed in PubMed for studies published between 14 November 2024 and 8 August 2025. Search strategies combined terms relating to Alzheimer’s disease (AD), mild cognitive impairment (MCI) and cognitively unimpaired (CU) groups; blood-based fluid biomarkers; and ethnicity, race or country income level. Full search strategies for all databases are provided in Supplementary Table 23.

### Eligibility criteria

Studies were eligible if they: (1) included at least one group with sporadic AD and/or MCI, diagnosed according to established clinical criteria, together with a CU comparison group, and/or stratified participants according to amyloid-β (Aβ) PET status (Aβ-positive versus Aβ-negative); (2) quantified biomarker concentrations in human plasma or serum using quantitative commercial assay platforms; (3) reported biomarker concentrations in pg/ml, or in formats convertible to pg/ml, as mean ± s.d., mean ± s.e.m., median with interquartile range (IQR), quartiles, or minimum and maximum values; and (4) reported at least one of the following biomarkers: Aβ40, Aβ42, Aβ42/Aβ40, p-tau181, p-tau217, p-tau231, t-tau, NfL or GFAP.

Studies were excluded if they: (1) included fewer than 10 participants in any comparison group; (2) used control groups with inflammatory, neurological or psychiatric conditions likely to influence biomarker concentrations; (3) measured biomarkers in cell cultures, tissue, animal models, cerebrospinal fluid or blood exosomes only; (4) used non-quantitative or semi-quantitative methods, or in-house assay platforms; (5) reported biomarker data only in non-convertible formats, such as z scores; or (6) were reviews, systematic reviews, meta-analyses, case reports, conference abstracts, book chapters, letters, commentaries or editorials. Only English-language publications were included.

### Study selection

Records were deduplicated and screened in Covidence (Veritas Health Innovation, Melbourne, Australia). Two reviewers (X.M. and G.H.) independently screened titles and abstracts, followed by full-text assessment of potentially eligible articles. Disagreements were resolved through discussion and, where required, consultation with two additional reviewers (A.P. and P.S.). The study selection process is shown in Fig. 1.

### Data extraction and data items

Data extraction was performed by one reviewer (X.M.) and checked by a second reviewer (G.H.), with uncertainties resolved through discussion with A.P. and P.S. Extracted data included: study identifiers; cohort name; publication year; continent and country or region; country income level; predominant cohort ethnoracial composition; demographic characteristics (age, sex and years of education); APOE ε4 carrier frequency; diagnostic criteria for AD and MCI; use of biomarker-supported diagnosis through PET, cerebrospinal fluid or autopsy; sample type (plasma or serum); biomarker concentrations; assay platform; sample size; and measurement units. For prospective studies, only baseline measurements were extracted. When multiple publications from the same cohort reported the same biomarker, the publication with the largest sample size was retained to avoid duplication. When a publication reported more than one cohort, cohorts were extracted separately. When multiple assay platforms were reported within a study, data from the most commonly used platform were extracted.

Where numerical data were presented only graphically, values were extracted using GetData Graph Digitizer (S. Fedorov, Russia). Corresponding authors were contacted when clarification was required regarding missing data or potential overlap between study populations.

### Population classification

Predominant cohort ethnoracial composition was classified at the study level as White, Black, Hispanic/Latino, Asian, MENA, or Indigenous when at least 80% of participants belonged to one group, based on study-reported participant characteristics, cohort descriptions or, where necessary, census information corresponding to the region and period of sample collection. Cohorts not meeting the 80% threshold were classified as multi-ethnic. For Asian cohorts, a more granular regional classification was also recorded (East Asian, Southeast Asian or South Asian). Country income level was classified using World Bank categories as high-income country (HIC) or low- and middle-income country (LMIC), according to the primary location of each cohort. The source of ethnoracial classification was additionally recorded as explicit when reported directly by the study, predominant when based on a predominantly single ethnoracial group (≥80%), or context-assigned when not explicitly reported and inferred from cohort context.

### Diagnostic approach

Diagnostic approach was categorized as biomarker-based when AD or MCI classification incorporated AD-related PET, cerebrospinal fluid biomarkers or autopsy confirmation. Diagnoses based solely on clinical criteria were categorized as clinical-based.

### Risk of bias assessment

Risk of bias was assessed independently by two reviewers (X.M. and G.H.) using a refined Newcastle–Ottawa Scale (NOS) adapted for this review (Supplementary Tables 24 and 25). When a single publication contributed more than one comparison (AD versus CU, MCI versus CU, or Aβ-positive versus Aβ-negative), each comparison was assessed separately (Supplementary Tables 26-28). Discrepancies were resolved by discussion, with arbitration by A.P. and P.S. when necessary.

### Data synthesis

The primary effect measure was the ratio of mean biomarker concentrations between comparison groups, expressed on the natural logarithmic scale. Biomarker concentrations were extracted separately for each case and comparison group for AD versus CU, MCI versus CU and Aβ-positive versus Aβ-negative analyses. When studies reported medians and IQRs, means and standard deviations were estimated using established methods^47-49^. When only medians and ranges were available, means and standard deviations were derived using published conversion approaches^47,48^. When s.e.m. was reported, standard deviations were calculated from the reported sample size. For log-transformed biomarker data, reported values were back-transformed to the original scale before analysis.

For each study, effect sizes were calculated as the natural logarithm of the ratio of mean biomarker concentrations between case and comparison groups. Standard errors were derived from the corresponding group means, standard deviations and sample sizes. Pooled estimates were then exponentiated and are presented as geometric mean ratios.

### Statistical analysis

Random-effects meta-analyses were used throughout. Between-study variance (τ^2^) was estimated using restricted maximum likelihood, and studies were weighted by the inverse of the squared standard error. Heterogeneity was assessed using Cochran’s Q statistic, I^2^ and τ^2 50^.

Primary meta-analyses were performed separately for AD versus CU, MCI versus CU and Aβ-positive versus Aβ-negative comparisons for each biomarker. Sensitivity analyses were performed by excluding studies with NOS scores <4, stratifying by assay platform, and restricting analyses to studies that included all three clinical groups (AD, MCI and CU), restricting analyses to studies using plasma samples, restricting analyses to cohorts with explicit or predominant ethnoracial classification, and performing leave-one-out analyses.

Subgroup meta-analyses stratified by predominant cohort ethnoracial composition were performed for each biomarker where at least two studies were available within a subgroup. Differences between population subgroups were assessed using random-effects meta-regression with subgroup as a categorical moderator. For clinically defined comparisons, additional models adjusted these contrasts for study-level age and sex where available; for Aβ-PET-defined comparisons, additional models adjusted for study-level age where available.

Exploratory interaction meta-regression analyses were then performed to examine whether study-level variation in biomarker ratios differed according to age, sex, diagnostic approach, country income level and geographic region. These analyses focused primarily on contrasts between predominantly Asian and predominantly White cohorts, with additional descriptive analyses for other population groups where sufficient data were available.

Small-study effects were assessed using funnel plots and Egger’s regression test, with trim-and-fill analyses applied where asymmetry was detected^51,52^. All statistical analyses and visualizations were conducted in R version 4.4.1 using the metafor and tidyverse packages^53,54^.

## Data Availability

All data used in this systematic review and meta-analysis were extracted from published articles, publicly available supplementary materials, and study-level information clarified by corresponding authors where required. The extracted study-level data supporting the analyses are provided in the Supplementary Tables. No individual-level participant data were used.

## Data availability

Extracted data, analysis datasets, data extraction templates and analytic code will be considered for sharing with researchers upon reasonable request to the corresponding author.

## Acknowledgements

We thank all participants and investigators of the original studies included in this meta-analysis. We also thank Helen Jones (UNSW Library) for assistance with the literature search strategy and database searching.

## Author contributions

X.M. and G.K.H. contributed to study design, literature screening, data extraction and data curation. X.M. performed the analyses, interpreted the results and wrote the manuscript. T.J. contributed to study design and interpretation of the findings. J.C. provided statistical input and contributed to interpretation of the analyses. P.S.S. and A.P. conceived and supervised the study, contributed to interpretation of the findings, and critically revised the manuscript. All authors reviewed the manuscript and approved the final version.

---

**Extended Data Fig. 1.**
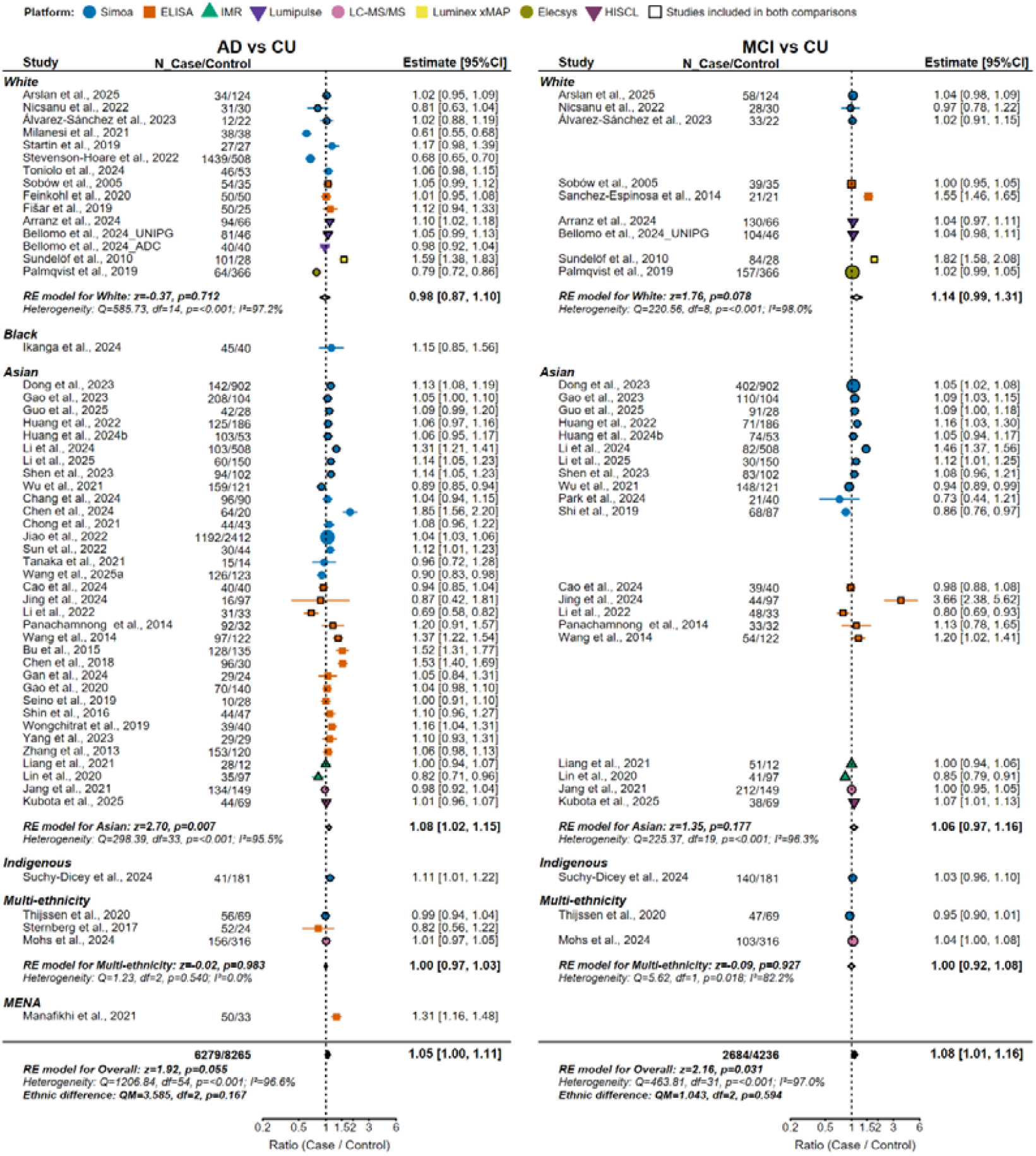
Forest plot of blood Aβ40 across ethnic groups. Forest plots showing the ratios of blood Aβ40 concentrations comparing AD with CU (left) and MCI with CU (right). Effect sizes are presented as geometric mean ratios (case⍰/⍰control) with 95% confidence intervals (CI). Study estimates are shown as points, with colors and shapes indicating assay platforms; markers outlined in black denote studies contributing to both comparisons. Hollow diamonds represent pooled estimates from random-effects (RE) models for each ethnic group, and filled diamonds represent the overall pooled effect. For each ethnic group and for the overall model, Cochran’s Q statistic, degrees of freedom (df), and I^2^ quantify between-study heterogeneity, while the z statistic and p value correspond to the test of the pooled effect. The between-ethnicity heterogeneity statistic (QM) is also reported for the overall model.

**Extended Data Fig. 2.**
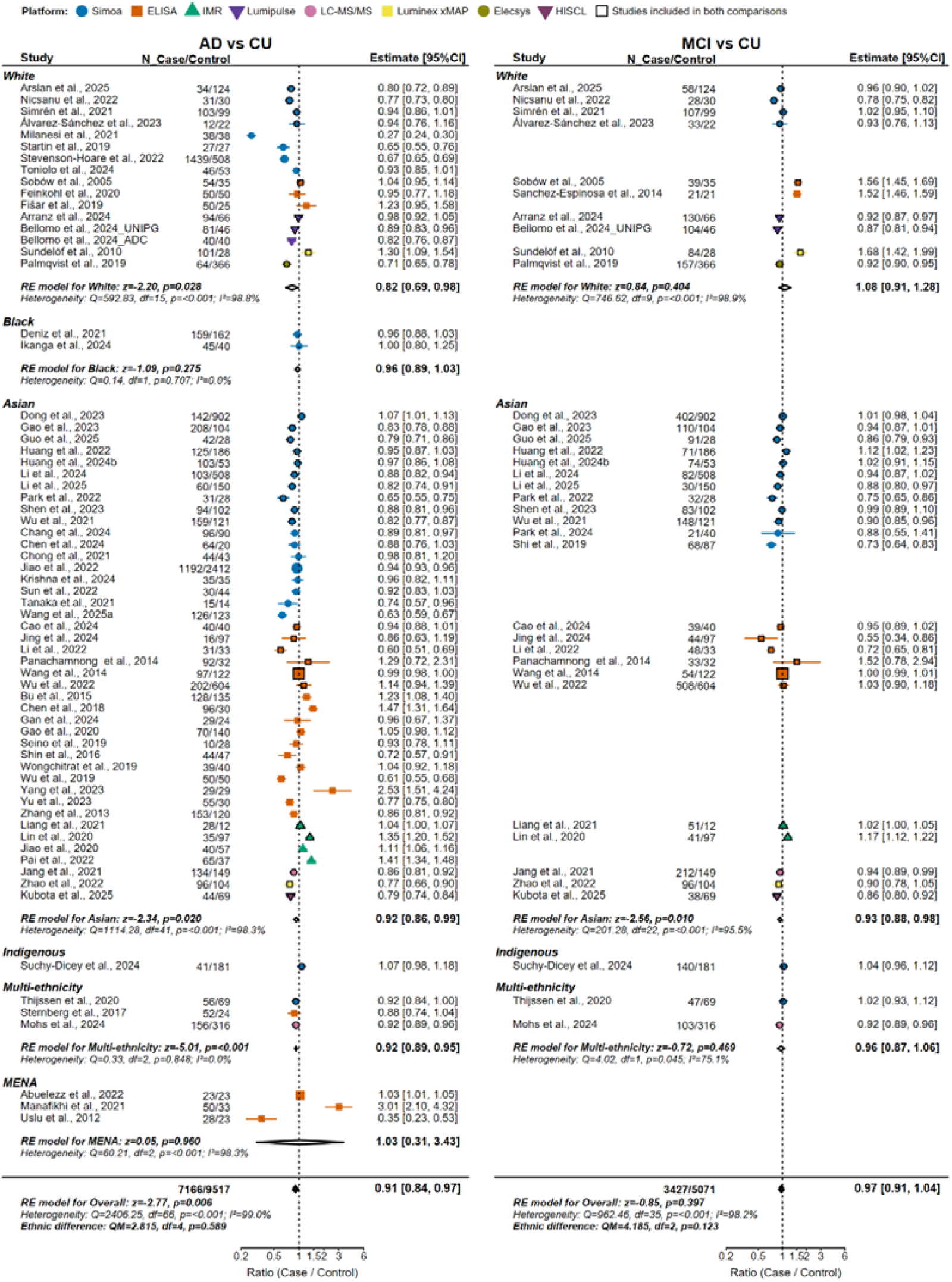
Forest plot of blood Aβ42 across groups defined by predominant cohort ethnoracial composition. Forest plots showing the ratios of blood Aβ42 concentrations comparing AD with CU (left) and MCI with CU (right). Effect sizes are presented as geometric mean ratios (case⍰/⍰control) with 95% confidence intervals (CI). Study estimates are shown as points, with colors and shapes indicating assay platforms; markers outlined in black denote studies contributing to both comparisons. Hollow diamonds represent pooled estimates from random-effects (RE) models for each ethnic group, and filled diamonds represent the overall pooled effect. For each ethnic group and for the overall model, Cochran’s Q statistic, degrees of freedom (df), and I^2^ quantify between-study heterogeneity, while the z statistic and p value correspond to the test of the pooled effect. The between-ethnicity heterogeneity statistic (QM) is also reported for the overall model.

**Extended Data Fig. 3.**
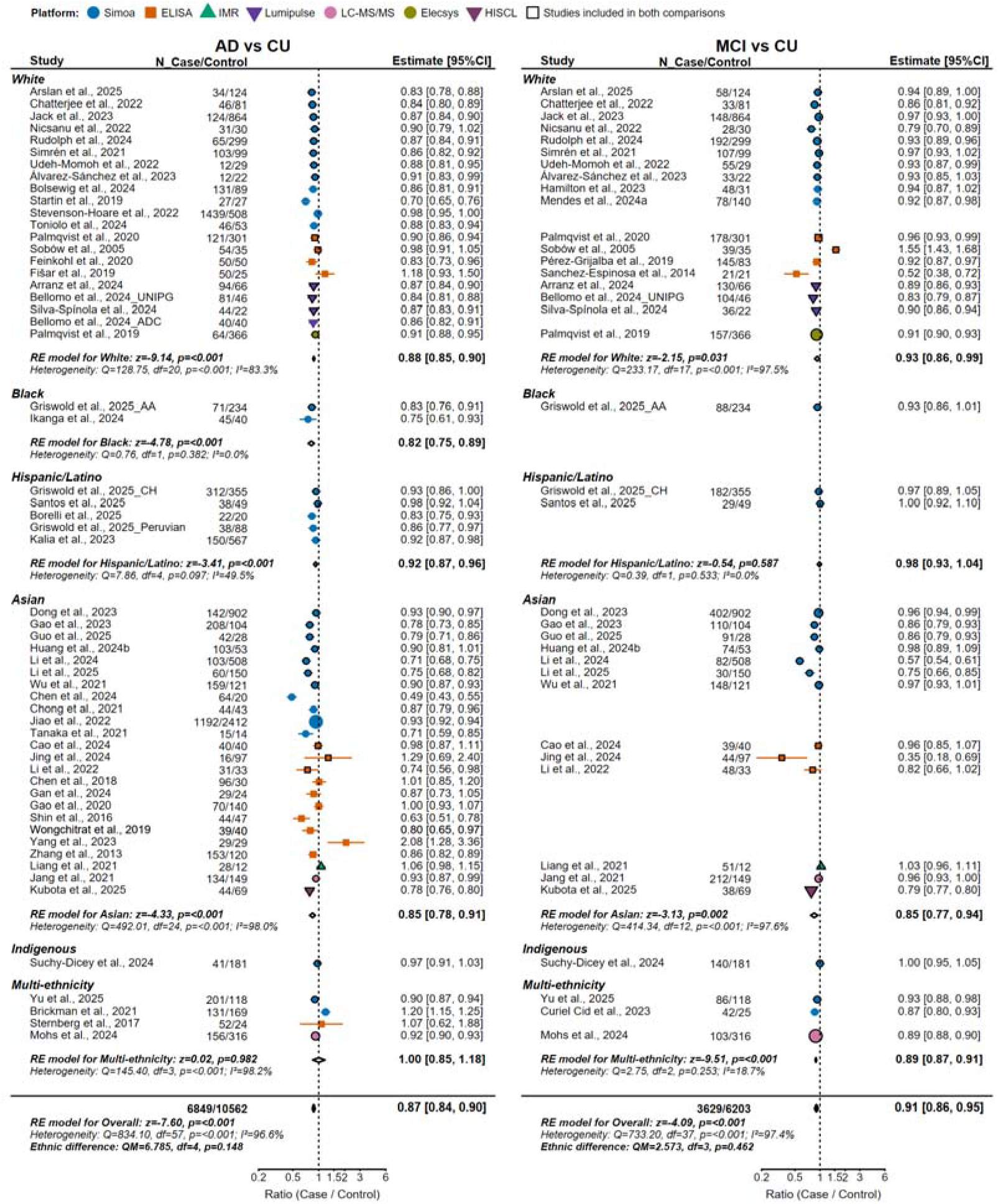
Forest plot of blood Aβ42/Aβ40 across groups defined by predominant cohort ethnoracial composition. Forest plots showing the ratios of blood Aβ42/Aβ40 comparing AD with CU (left) and MCI with CU (right). Effect sizes are presented as geometric mean ratios (case⍰/ ⍰control) with 95% confidence intervals (CI). Study estimates are shown as points, with colors and shapes indicating assay platforms; markers outlined in black denote studies contributing to both comparisons. Hollow diamonds represent pooled estimates from random-effects (RE) models for each ethnic group, and filled diamonds represent the overall pooled effect. For each ethnic group and for the overall model, Cochran’s Q statistic, degrees of freedom (df), and I^2^ quantify between-study heterogeneity, while the z statistic and p value correspond to the test of the pooled effect. The between-ethnicity heterogeneity statistic (QM) is also reported for the overall model.

**Extended Data Fig. 4.**
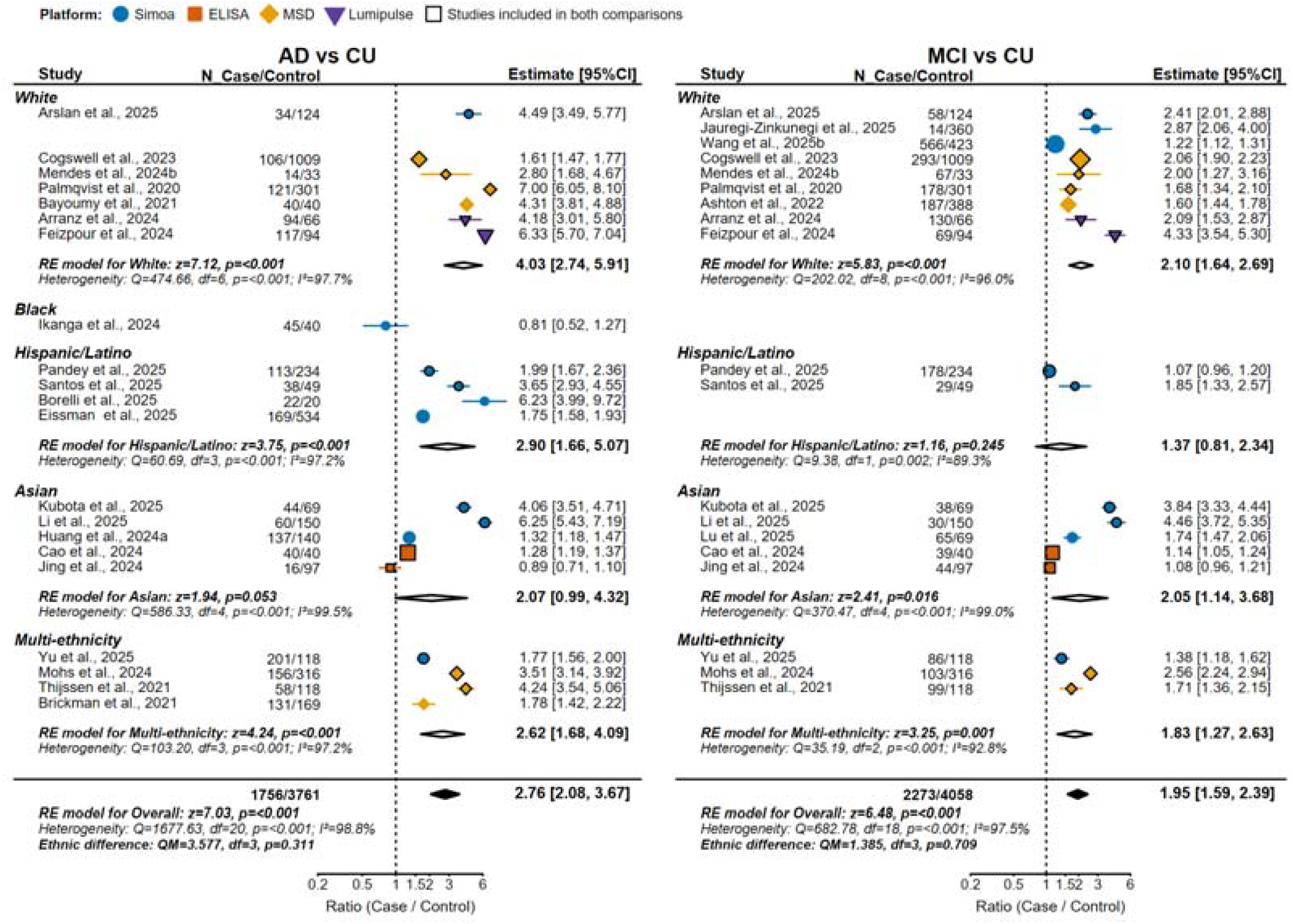
Forest plot of blood p-tau217 across groups defined by predominant cohort ethnoracial composition. Forest plots showing the ratios of blood p-tau217 concentrations comparing AD with CU (left) and MCI with CU (right). Effect sizes are presented as geometric mean ratios (case⍰/ ⍰control) with 95% confidence intervals (CI). Study estimates are shown as points, with colors and shapes indicating assay platforms; markers outlined in black denote studies contributing to both comparisons. Hollow diamonds represent pooled estimates from random-effects (RE) models for each ethnic group, and filled diamonds represent the overall pooled effect. For each ethnic group and for the overall model, Cochran’s Q statistic, degrees of freedom (df), and I^2^ quantify between-study heterogeneity, while the z statistic and p value correspond to the test of the pooled effect. The between-ethnicity heterogeneity statistic (QM) is also reported for the overall model.

**Extended Data Fig. 5.**
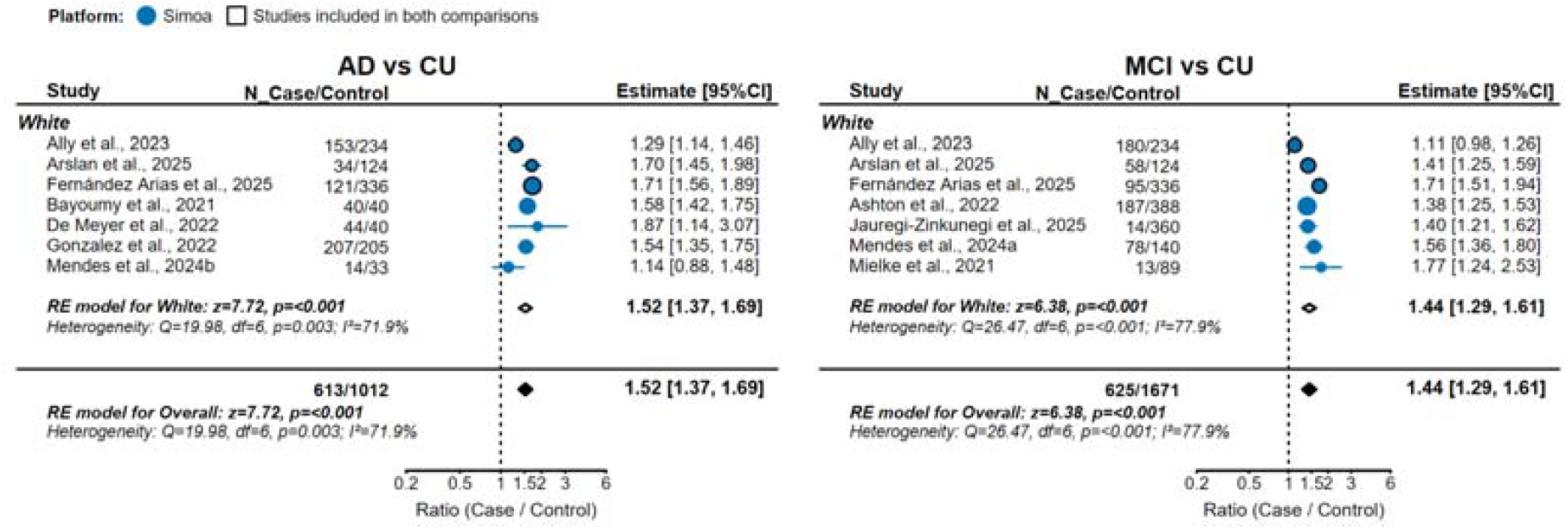
Forest plot of blood p-tau231 across groups defined by predominant cohort ethnoracial composition. Forest plots showing the ratios of blood p-tau231 concentrations comparing AD with CU (left) and MCI with CU (right). Effect sizes are presented as geometric mean ratios (case⍰/ ⍰control) with 95% confidence intervals (CI). Study estimates are shown as points, with colors and shapes indicating assay platforms; markers outlined in black denote studies contributing to both comparisons. Hollow diamonds represent pooled estimates from random-effects (RE) models for each ethnic group, and filled diamonds represent the overall pooled effect. For each ethnic group and for the overall model, Cochran’s Q statistic, degrees of freedom (df), and I^2^ quantify between-study heterogeneity, while the z statistic and p value correspond to the test of the pooled effect. The between-ethnicity heterogeneity statistic (QM) is also reported for the overall model.

**Extended Data Fig. 6.**
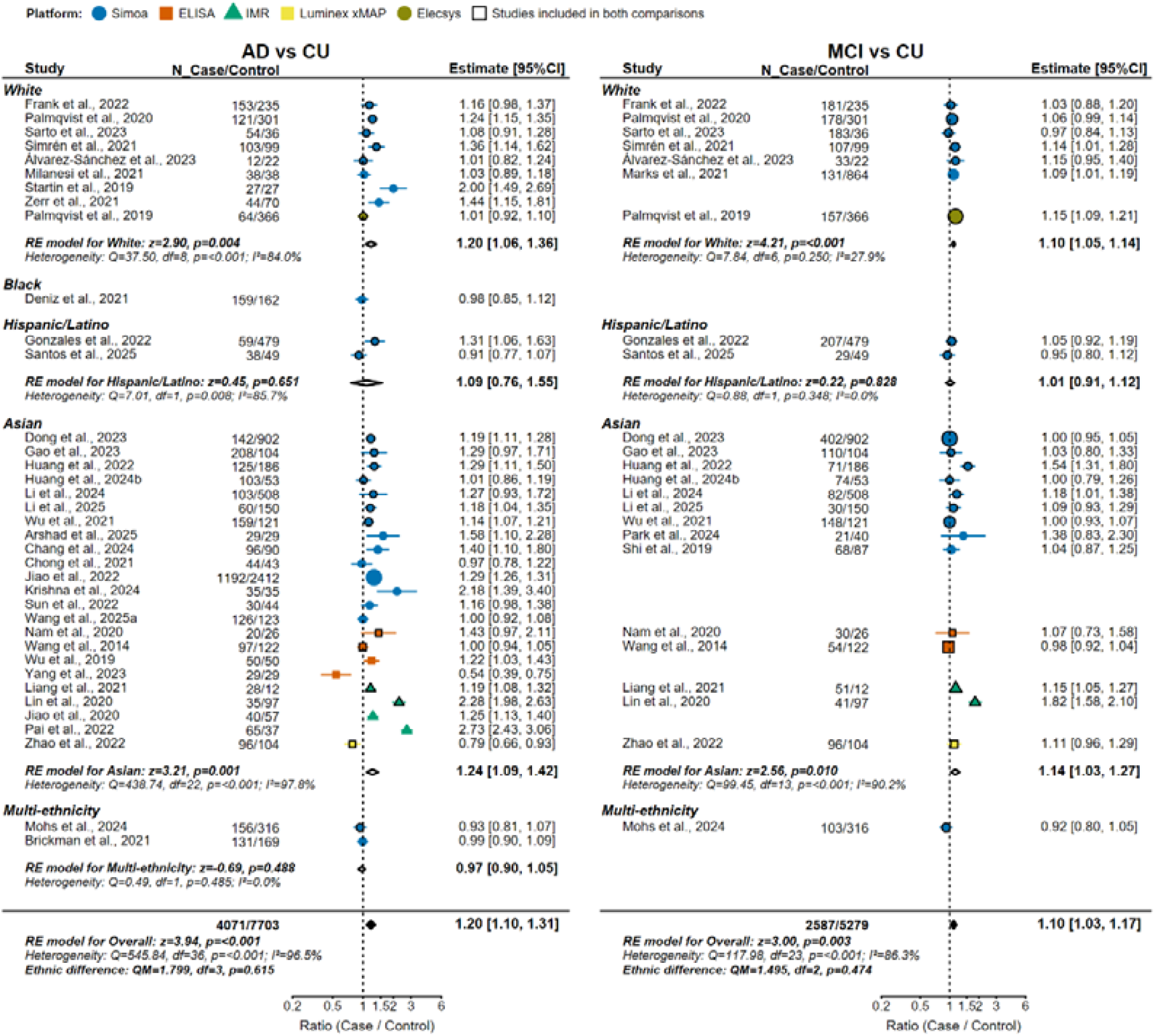
Forest plot of blood t-tau across groups defined by predominant cohort ethnoracial composition. Forest plots showing the ratios of blood t-tau concentrations comparing AD with CU (left) and MCI with CU (right). Effect sizes are presented as geometric mean ratios (case⍰/ ⍰control) with 95% confidence intervals (CI). Study estimates are shown as points, with colors and shapes indicating assay platforms; markers outlined in black denote studies contributing to both comparisons. Hollow diamonds represent pooled estimates from random-effects (RE) models for each ethnic group, and filled diamonds represent the overall pooled effect. For each ethnic group and for the overall model, Cochran’s Q statistic, degrees of freedom (df), and I^2^ quantify between-study heterogeneity, while the z statistic and p value correspond to the test of the pooled effect. The between-ethnicity heterogeneity statistic (QM) is also reported for the overall model.

**Extended Data Fig. 7.**
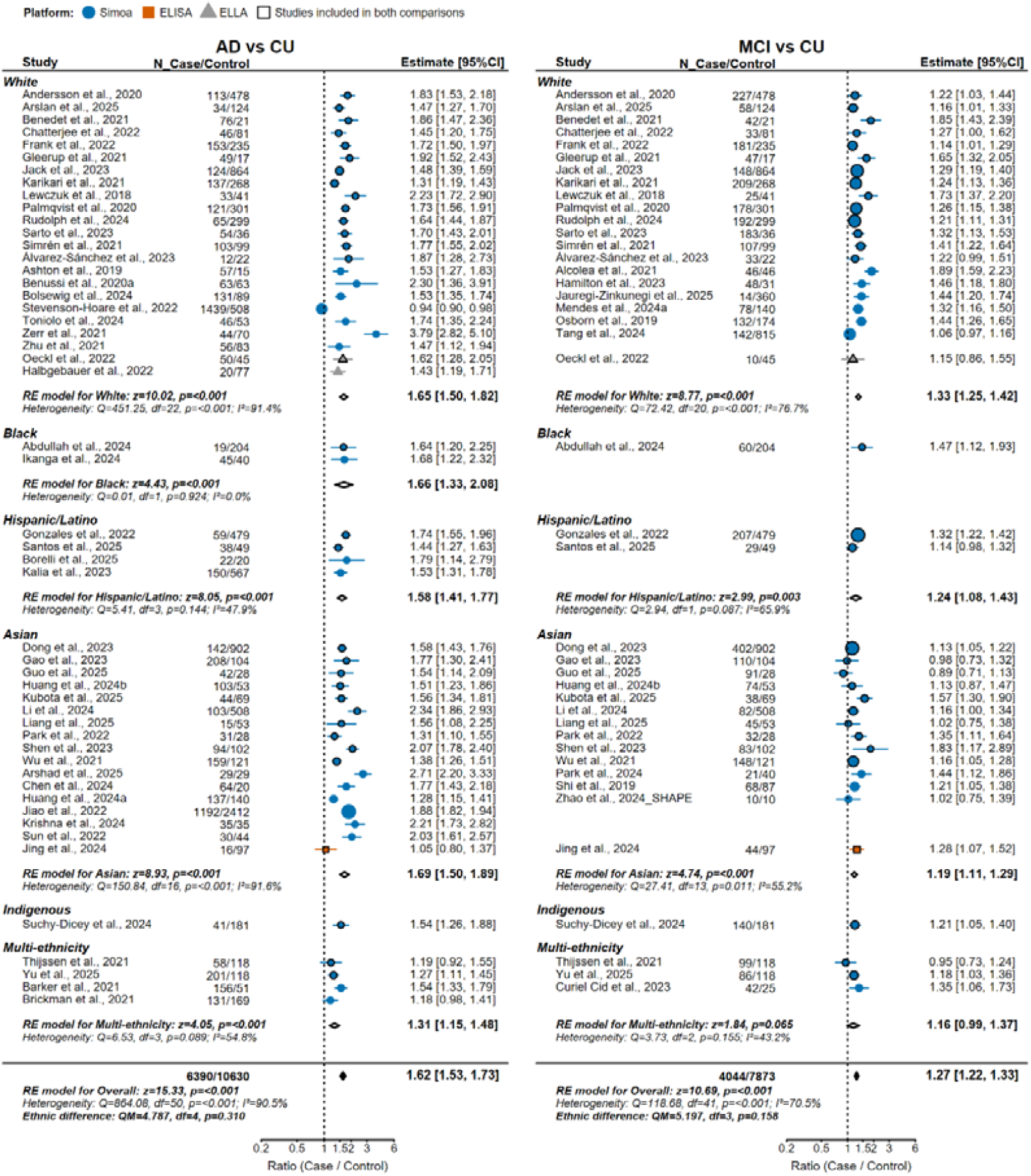
Forest plot of blood NfL across groups defined by predominant cohort ethnoracial composition. Forest plots showing the ratios of blood NfL concentrations comparing AD with CU (left) and MCI with CU (right). Effect sizes are presented as geometric mean ratios (case⍰/⍰control) with 95% confidence intervals (CI). Study estimates are shown as points, with colors and shapes indicating assay platforms; markers outlined in black denote studies contributing to both comparisons. Hollow diamonds represent pooled estimates from random-effects (RE) models for each ethnic group, and filled diamonds represent the overall pooled effect. For each ethnic group and for the overall model, Cochran’s Q statistic, degrees of freedom (df), and I^2^ quantify between-study heterogeneity, while the z statistic and p value correspond to the test of the pooled effect. The between-ethnicity heterogeneity statistic (QM) is also reported for the overall model.

**Extended Data Fig. 8.**
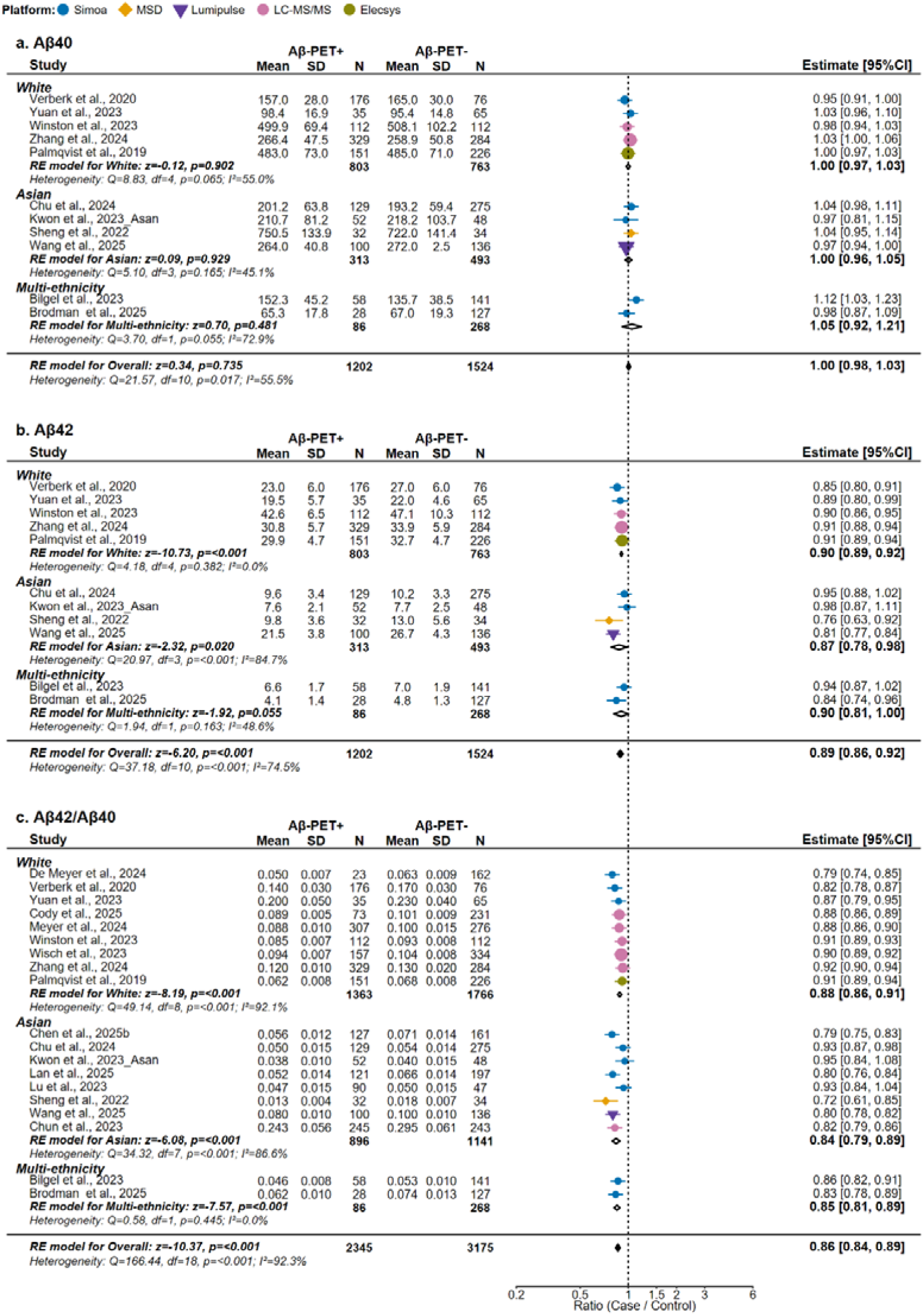
Forest plots of β-amyloid-related blood biomarkers in Aβ-PET-positive vs Aβ-PET-negative individuals across groups defined by predominant cohort ethnoracial composition. Forest plots showing blood Aβ40 (a), Aβ42 (b), and Aβ42/Aβ40 (c) comparing Aβ-PET-positive and Aβ-PET-negative individuals across groups defined by predominant cohort ethnoracial composition. Effect sizes are expressed as geometric mean ratios (Aβ-PET+⍰/ ⍰Aβ-PET-) with 95% confidence intervals (CI). Study estimates are shown as points, with colors and shapes indicating assay platforms. Hollow diamonds indicate pooled estimates from random-effects (RE) models for each ethnic group, and filled diamonds indicate the overall pooled effect. For each ethnic group and for the overall model, Cochran’s Q statistic, degrees of freedom (df), and I^2^ quantify between-study heterogeneity, while the z statistic and p value correspond to the test of the pooled effect. The between-ethnicity heterogeneity statistic (QM) is also reported for the overall model.

**Extended Data Fig. 9.**
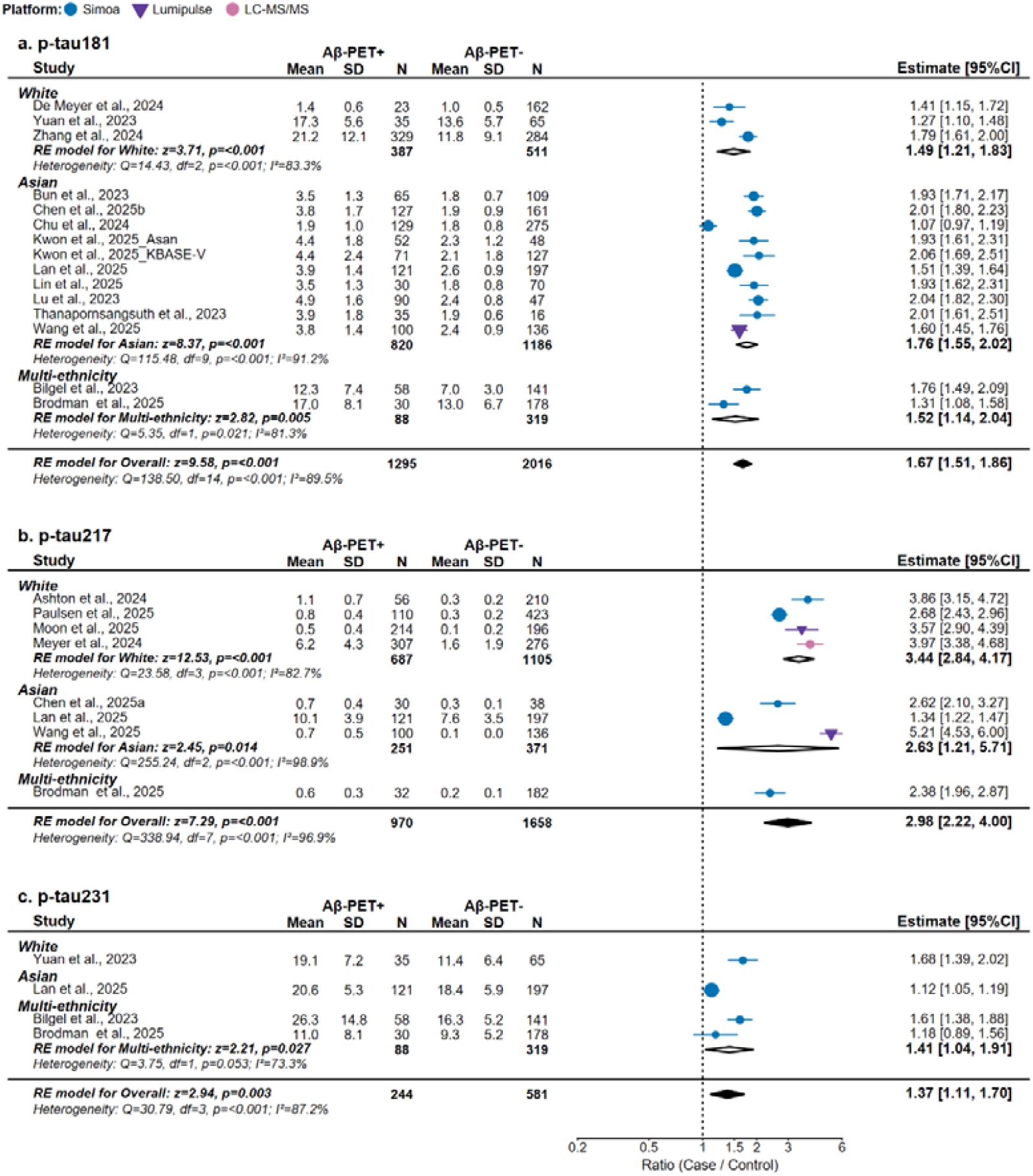
Forest plots of phosphorylated tau blood biomarkers in Aβ-PET-positive vs Aβ-PET-negative individuals across groups defined by predominant cohort ethnoracial composition. Forest plots showing blood p-tau181 (a), p-tau217 (b), and p-tau231 (c) comparing Aβ-PET-positive and Aβ-PET-negative individuals across groups defined by predominant cohort ethnoracial composition. Effect sizes are expressed as geometric mean ratios (Aβ-PET+⍰/ ⍰Aβ-PET-) with 95% confidence intervals (CI). Study estimates are shown as points, with colors and shapes indicating assay platforms. Hollow diamonds indicate pooled estimates from random-effects (RE) models for each ethnic group, and filled diamonds indicate the overall pooled effect. For each ethnic group and for the overall model, Cochran’s Q statistic, degrees of freedom (df), and I^2^ quantify between-study heterogeneity, while the z statistic and p value correspond to the test of the pooled effect. The between-ethnicity heterogeneity statistic (QM) is also reported for the overall model.

**Extended Data Fig. 10.**
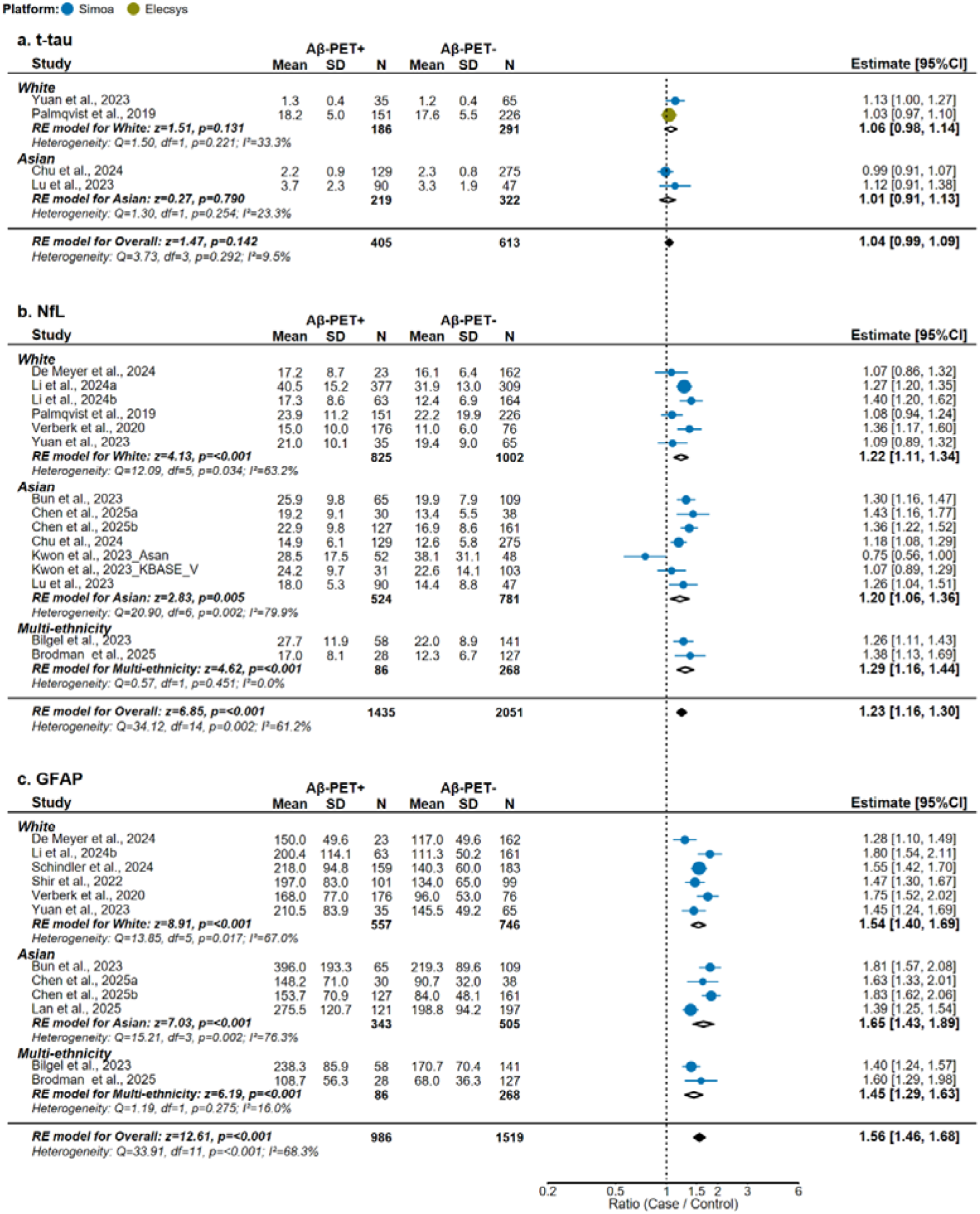
Forest plots of blood biomarkers of neurodegeneration and glial activation in Aβ-PET-positive vs Aβ-PET-negative individuals across groups defined by predominant cohort ethnoracial composition. Forest plots showing blood t-tau (a), NfL (b), and GFAP (c) comparing Aβ-PET-positive and Aβ-PET-negative individuals across ethnic groups. Effect sizes are expressed as geometric mean ratios (Aβ-PET+⍰/ ⍰Aβ-PET-) with 95% confidence intervals (CI). Study estimates are shown as points, with colors and shapes indicating assay platforms. Hollow diamonds indicate pooled estimates from random-effects (RE) models for each ethnic group, and filled diamonds indicate the overall pooled effect. For each ethnic group and for the overall model, Cochran’s Q statistic, degrees of freedom (df), and I^2^ quantify between-study heterogeneity, while the z statistic and p value correspond to the test of the pooled effect. The between-ethnicity heterogeneity statistic (QM) is also reported for the overall model.

