## Supplementary Figure 1 for "Blood biomarkers for Alzheimer disease across ethnoracial groups and healthcare settings: a systematic review and meta-analysis"

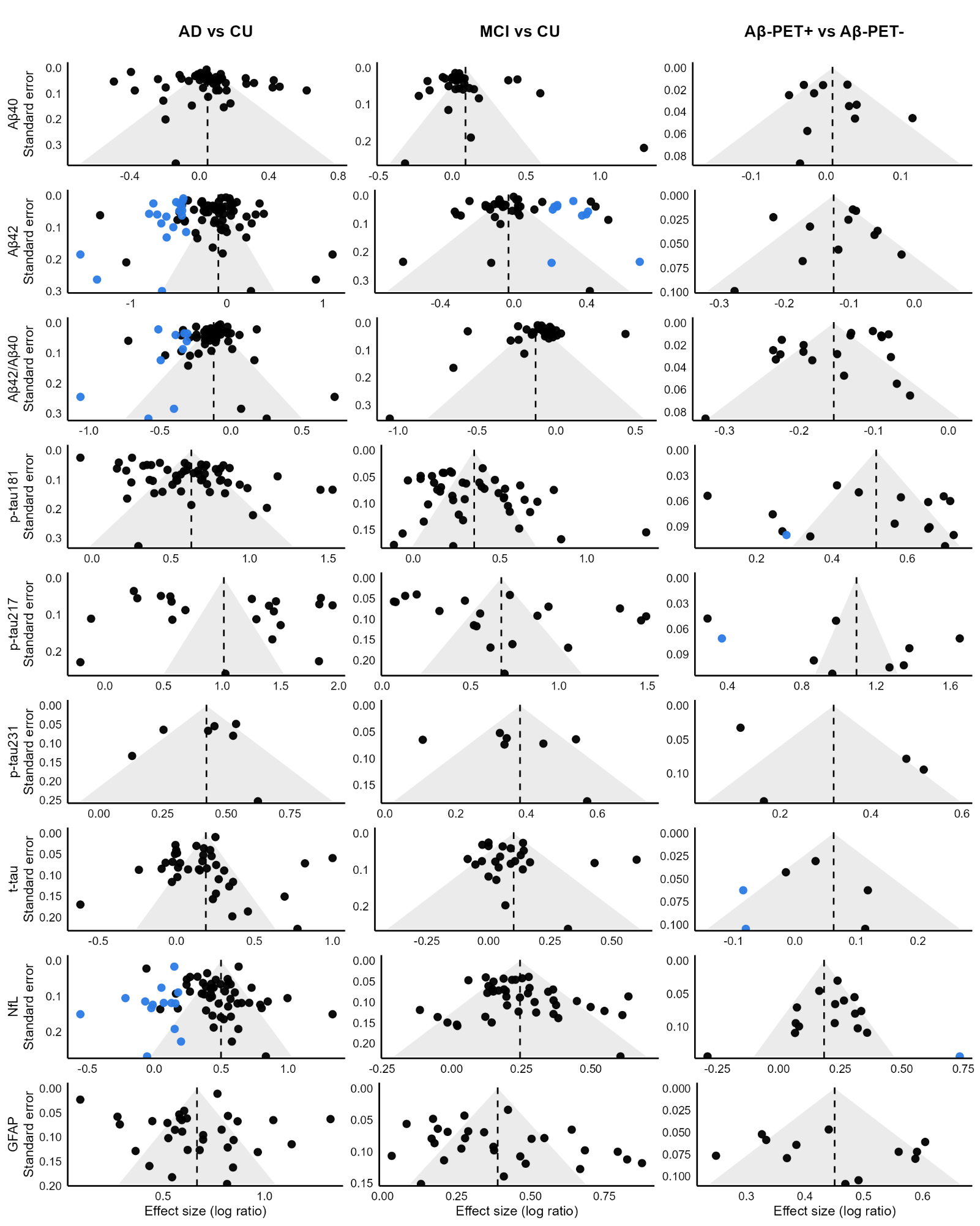

**Supplementary Fig. 1. Funnel plots assessing publication bias across biomarkers.** Funnel plots for nine blood biomarkers across three comparisons (AD vs CU, MCI vs CU, and Aβ-PET+ vs Aβ-PET-). Shaded region: expected 95% confidence bounds in the absence of small-study effects. Black points: observed studies. Blue points: studies identified or imputed during publication-bias diagnostics. Vertical dashed lines: pooled effect estimate for each comparison.
